# Molecular, haemodynamic, and functional effects of LSD in the human brain

**DOI:** 10.1101/2025.06.17.25329677

**Authors:** Drummond E-Wen McCulloch, Kristian Larsen, Annette Johansen, Kristian H. Reveles Jensen, Charlotte Havelund Nykjær, Friederike Holze, Nora Falck, Victor Neufeld, Emilia Steenstrup, Peter Skov-Andersen, Anders Spanggård, Maria Geisler, Paw Patrick Randrup, Peter Steen Jensen, Vladimir Shalgunov, Sys Stybe Johansen, Marie Katrine Klose Nielsen, Thomas Lund Andersen, Dea Siggaard Stenbæk, Claus Svarer, Patrick MacDonald Fisher, Gitte Moos Knudsen

**Author notes:** Shared first authorship (Drummond E-Wen McCulloch, Kristian Larsen). Shared senior authorship (Patrick MacDonald Fisher, Gitte Moos Knudsen). Correspondence (P.M.F.) or (G.M.K).

## Abstract

In this study, we provide the first study to integrate molecular and functional neuroimaging during psychedelic drug effects in humans. Using simultaneous PET-MRI technology, we describe multiple brain actions of lysergic acid diethylamide (LSD) in seven healthy volunteers blinded to LSD dose received. We quantify the occupancy of LSD at cerebral serotonin 2A receptors and show that LSD increases global cerebral blood flow (CBF) and internal carotid artery flow without affecting the diameter of the internal carotid artery, opposite effects to those observed following psilocybin. Functional connectivity analyses show decreases in global connectivity (GCOR). Change in GCOR is negatively correlated with change in CBF. We observe an anticlockwise hysteresis loop between plasma drug levels and subjective effects, suggesting atypical pharmacodynamic mechanisms. We contrast our CBF and GCOR findings and their relation in a separate cohort of 25 participants administered psilocybin, highlighting key consistencies and differences in their neural effects. By establishing the dose-occupancy relation of LSD in humans, our findings provide critical insights for the clinical development of psychedelic compounds and demonstrate unique neurophysiological effects that distinguish LSD from related psychedelics.

## Main

Lysergic acid diethylamide (LSD) is a serotonergic psychedelic drug (1). Recent phase 1 and 2a studies have demonstrated that doses of up to 200 μg LSD are safe (2–4), and that a single dose of LSD is associated with symptom reduction in patients with anxiety disorders, up to several months after treatment (5,6). LSD has recently entered three phase 3 trials for the treatment of anxiety and depressive disorders (NCT06941844, NCT06809595, NCT06741228). Intake of LSD changes mood, cognition, and perception from doses as low as 10 µg; the psychoactive effects increase dose-dependently, eliciting extremely intense effects at 100 µg and above (2,4,7). The promising clinical effects highlight the importance of understanding LSD’s mechanisms of action in the human brain.

The serotonin 2A receptor (5-HT2AR) is highly expressed in the human neocortex (8) and is the critical target for psychedelic effects, with agonist affinity correlating with potency in inducing subjective effects across psychedelics (9). This is supported by studies showing that ketanserin, a 5-HT2A/2CR antagonist, can block LSD’s psychedelic effects (3,4,10), and that selective 5-HT2AR agonists can produce psychedelic effects (11,12). Together, this shows that 5-HT2AR agonism is both necessary and sufficient for the psychedelic effects of LSD. Compared to psilocybin, which produces similar behavioural effects at appropriate doses (2), LSD shows distinct pharmacodynamics, with a reported delay between peak plasma levels and peak subjective effects of up to 1 hour (4,13). A potential explanation for this intriguing observation is based on in vitro crystallography data demonstrating that the binding pocket of LSD requires a conformational change in extracellular loops, which may result in very slow binding and dissociation between LSD and the receptor, the so-called “lid hypothesis” (14,15).

Understanding LSD’s neural effects in humans remains incomplete and has so far mainly been based on blood-oxygen level dependent (BOLD) functional magnetic resonance imaging (fMRI) studies (16). These have identified alterations in global and thalamocortical connectivity, and increases in a range of information-entropy metrics (17), but there has been extremely little replication in the space, and no investigations to date have evaluated relations to target engagement (18). Additionally, the role of the cerebral vasculature, which richly expresses vasoconstriction-mediating 5-HT2AR on vascular smooth muscle cells (19), has largely been neglected. Addressing these gaps around LSD’s receptor occupancy and associated systems-level brain function improves our ability to link molecular mechanisms to neural dynamics and therapeutic outcomes.

Leveraging simultaneous PET-MRI technology in healthy volunteers taking an oral dose of LSD, we report for the first time how plasma LSD is associated with in vivo cerebral 5-HT2AR occupancy and subjective drug effects. We present the first comprehensive evaluation of psychedelic effects on brain haemodynamics, revealing that LSD substantially increases internal carotid artery (ICA) flow velocity and cerebral blood flow (CBF), without affecting the ICA diameter. Moreover, when we quantify LSD effects on functional brain activity, we provide evidence for widespread decreases in global connectivity (GCOR) that are negatively related to changes in CBF. To add further context, we evaluate the relation between change in GCOR and CBF in an independent sample of 28 healthy volunteers following psilocybin administration, showing a similar negative relation. These complementary measures provide valuable insights into LSD’s neural effects that challenge some widely held beliefs about the acute neural effects of psychedelics.

## Results

### Baseline multimodal neuroimaging of participants

Eleven healthy participants (2 female; age: mean ± SD, 31 ± 9 years; weight: 81 ± 12 kg) underwent a multimodal baseline imaging session. See Supplementary Table S1 for demographic details. These included a [^11^C]Cimbi-36 PET scan to quantify neocortex 5-HT2AR nondisplaceable binding potential (BP_ND_), multi-band multi-echo fMRI to assess brain activity, and haemodynamic measurements combining ICA angiography, flow quantification with phase contrast mapping (PCM), and CBF measurement with pseudo-continuous arterial spin labelling (pcASL). Baseline 5-HT2AR BP_ND_ showed high binding in the neocortex (mean±SD = 1.11±0.26). Participants listened to a predefined playlist during the entire scan to increase comfort (see Methods for more details). The preprocessed fMRI data exhibited modular network organisation characteristic of healthy brain function (20), with normalised modularity values averaging 3.65±0.62 (mean±SD) times higher than those observed in random networks, confirming community structure in the brain’s connectome. CBF estimates (mean±SD = 49.2±10.4 ml/100g/min) (21), ICA flow (mean±SD = 215.0±57.8 ml/min), and ICA diameter (mean±SD = 4.65±0.32 mm, see Table S2) were all within expected ranges (22–24). LSD is metabolised by the CYP2D6 enzyme, which has genetic variations that affect enzymatic function (25). Two participants were intermediate metabolisers (n = 1 with activity score 0.5 and n = 1 with score 1.0), and the remaining nine participants were normal metabolisers (n = 4 with score 1.5, n = 5 with score 2.0). The observed relation between peak plasma LSD level (PLL) and dose administered was not clearly modified by *CYP2D6* genotype (Supplementary Figures S1, S2).

### LSD pharmacokinetics and acute subjective effects

On a separate day, participants were administered a single oral dose of LSD tartrate (25-200 μg freebase equivalent). See Table 1 for an overview of all occupancies, plasma concentrations, and doses administered at each scan. This dose range was evaluated to estimate the relation to 5-HT2AR occupancy across a clinically relevant range of dose and PLL. In a dose-dependent manner, LSD intake produced psychedelic effects in all participants, without serious adverse effects (Supplementary results). PLL peaked after 96 mins, and subjective drug intensity (SDI) peaked 30 mins later (Figure 1A, B). A clear anticlockwise hysteresis loop emerged when examining the relation between plasma LSD levels and SDI scores over time (Figure 1C) such that lower SDI scores are reported during the ascending phase compared to the descending phase at equivalent PLL. This indicates that for a given PLL, subjective effects were weaker while plasma levels were rising than while they were falling. We did not observe this effect for psilocybin (n = 28) (Supplementary Figure S14). Seven participants completed PET-MRI scans during peak drug effects, and two completed scans after subjective drug effects had largely subsided. Despite scan sessions lasting over two hours each, all but one participant tolerated the procedure well, reporting feeling calmer, less tired, and less physically uncomfortable and more emotionally responsive to music than during the baseline scan (Supplementary Figure S3, S13). Only one participant (P10, 150 μg) experienced scanner-related anxiety that necessitated early scan termination; cessation of scanning resolved the anxiety. One participant (P9, 150 μg) experienced severe nausea shortly after drug administration and was not scanned while in the psychedelic state. For two participants (P8, 150 μg and P11, 75 μg), the scanner malfunctioned, and all scan data was lost.

**Table 1.** | Pharmacokinetic and receptor occupancy measurements across participants. Overview of LSD doses, peak plasma concentrations (Cmax), timing, and receptor occupancy data from PET scans. The table shows data for each participant including LSD dose (µg, freebase equivalent), maximum observed plasma LSD concentration (nM), timing of PET scans relative to LSD administration (minutes), mean plasma LSD concentration during scan windows (nM), mean subjective drug intensity (SDI) ratings during scans, and measured 5-HT2AR occupancy (%). Data from both peak and late scans are shown where assessed. P8-11 had only baseline PET data collected. SDI was rated on a 0-10 scale, where 0 represents no drug effects and 10 represents the strongest imaginable effects. Receptor occupancy was calculated using [^11^C]Cimbi-36 PET. Time post-LSD refers to the minutes between LSD administration and radiotracer injection.

|  | LSD dose | Cmax | Peak Scan |  |  |  | Late Scan |  |  |  |
| --- | --- | --- | --- | --- | --- | --- | --- | --- | --- | --- |
|  |  |  | Time post-dose | [LSD] | SDI | Occupancy | Time post-dose | [LSD] | SDI | Occupancy |
|  | µg | nM | min | nM | a.u. |  | min | nM | a.u. |  |
| 1 | 25 | 1.37 | 175 | 1.23 | 3.0 | 48% |  |  |  |  |
| 2 | 50 | 4.08 | 164 | 3.32 | 10.0 | 42% |  |  |  |  |
| 3 | 75 | 8.77 | 175 | 7.22 | 7.5 | 75% |  |  |  |  |
| 4 | 75 | 9.21 | 161 | 6.43 | 8.5 | 85% | 420 | 3.84 | 1.5 | 74% |
| 5 | 100 | 9.56 | 271 | 5.31 | 8.0 | 67% | 522 | 2.25 | 1.0 | 12% |
| 6 | 100 | 6.19 | 167 | 6.19 | 9.0 | 73% |  |  |  |  |
| 7 | 200 | 8.52 | 168 | 8.08 | 10.0 | 83% |  |  |  |  |
| 8 | 150 | 15.26 |  |  |  |  |  |  |  |  |
| 9 | 150 | 7.30 |  |  |  |  |  |  |  |  |
| 10 | 150 | 5.67 |  |  |  |  |  |  |  |  |
| 11 | 75 | 3.67 |  |  |  |  |  |  |  |  |

**Figure 1.**
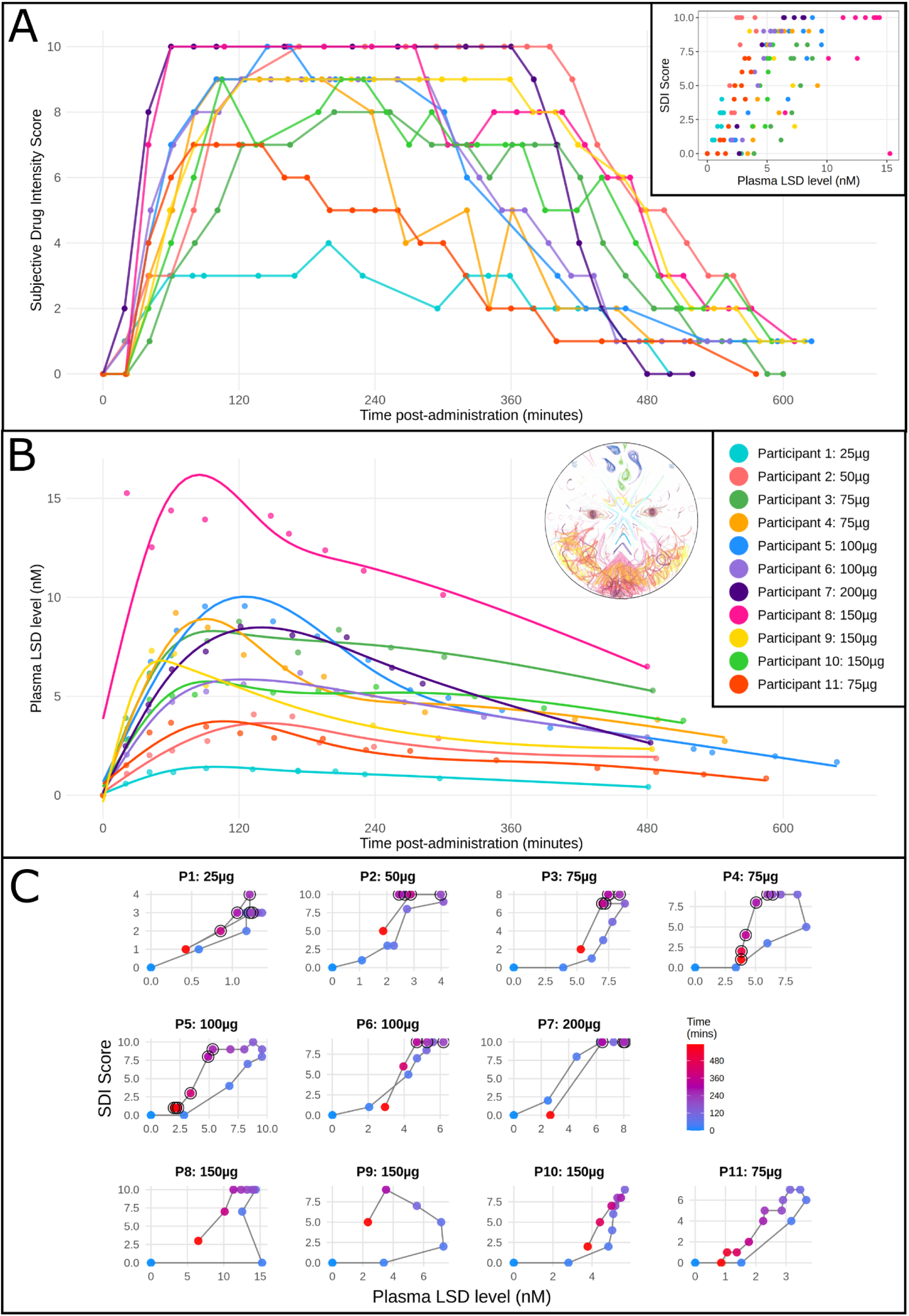
| Subjective and pharmacokinetic effects of LSD intake. A, Time course of subjective drug intensity (SDI) scores (0–10) for each participant following LSD administration (25-200 µg); insert shows SDI plotted against plasma LSD concentration. B, Plasma LSD concentrations over time post LSD (intake is t=0). Curves are fitted with a spline for visualisation only. Subplot includes a representative drawing of the LSD experience by P8 (pink curve). C, Single-subject SDI vs PLL hysteresis plots. Points are chronologically connected by black lines, and points are coloured by time, i.e., the number of minutes post-LSD intake. Black outline circles indicate timepoints during PET scans. P4 and P5 received two PET scans during LSD effects. All but P7 show an anticlockwise hysteresis loop. A similar plot to C, but for psilocybin, is available in Supplementary Figure S14 and does not show hysteresis effects.

### Occupancy of LSD at the 5-HT2AR is associated with subjective drug effects

By comparing [^11^C]Cimbi-36 PET at baseline and after LSD administration (Figure 2A), we calculated neocortical 5-HT2AR LSD occupancy. We fit the occupancy data and PLL measured during the scan to a Hill-Langmuir model (R^2^ = 0.60) with bootstrapped confidence intervals. This revealed an IC_50_ of 1.93 nM (95% CI: 0.53-2.79 nM) equivalent to 0.62 ng/ml (CI: 0.17-0.9 ng/ml) and maximum occupancy (Occ_max_) of 97.4% (95% CI: 75.3-100%) (Figure 2B). A similar relation was observed between LSD dose and occupancy (R^2^ = 0.58), with an ED_50_ of 31.1 µg (95% CI: 7.6-46.4 µg) and Occ_max_ of 96.2% (95% CI: 73.5-100%) (Supplementary Figure 4A). When analysing body weight-adjusted LSD dose, we observed a closer relation with occupancy (R^2^ = 0.75), with an ED_50_ of 0.41 µg/kg (95% CI: 0.17-0.54 µg/kg) and Occ_max_ of 98.7% (95% CI: 80.3-100%) (Supplementary Figure 4B). Peak occupancy was related to mean within-scan SDI ratings with a sigmoidal fit (Figure 2C, R^2^ = 0.87).

**Figure 2.**
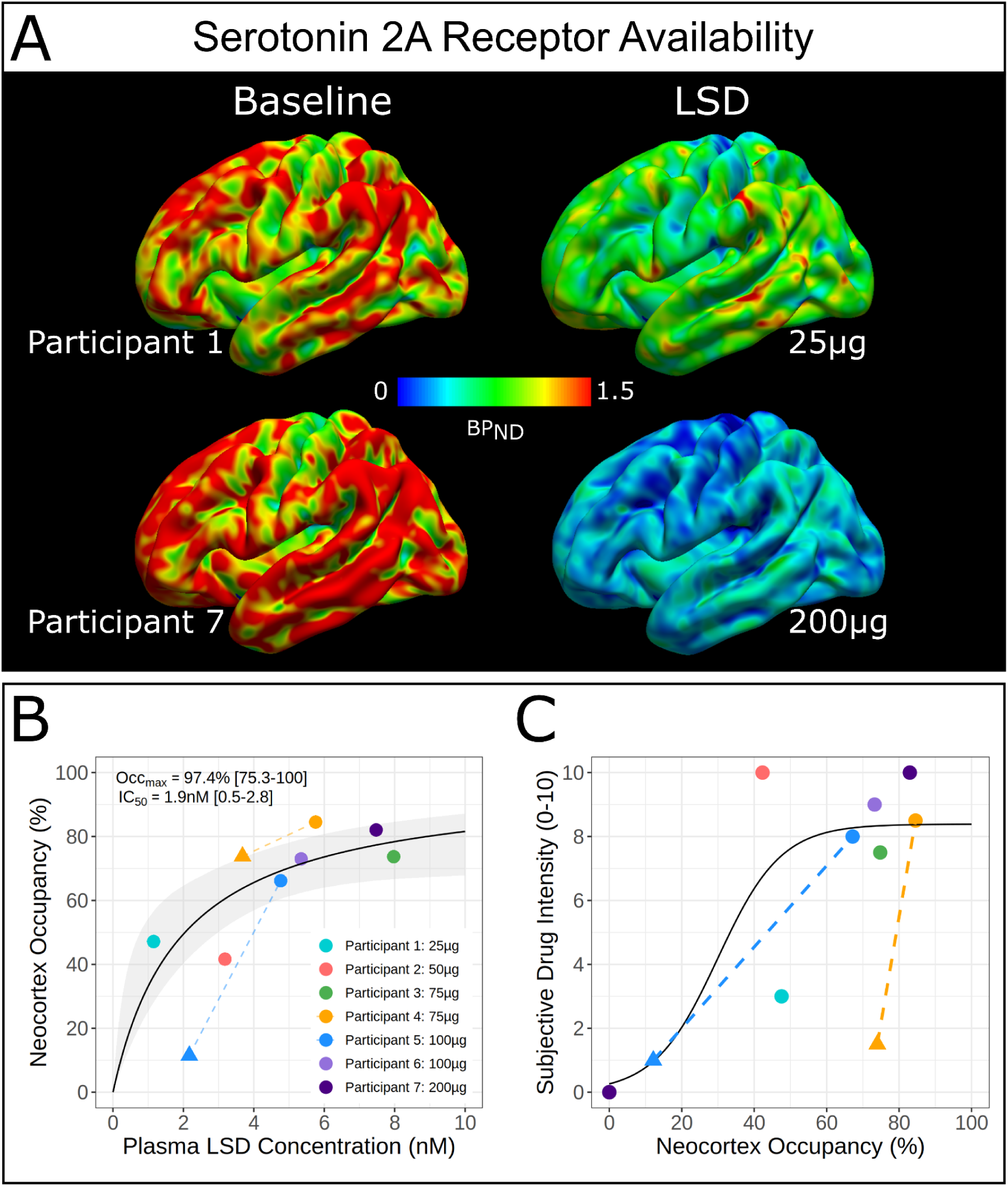
| Serotonin 2A receptor occupancy by LSD measured with [^11^C]Cimbi-36 PET. A, Cortical 5-HT2AR nondisplaceable binding potential (BP_ND_) maps at baseline and peak LSD for participants receiving 25 µg (top) and 200 µg (bottom) doses. The colour scale indicates BP_ND_ values. B, Plasma LSD concentration versus receptor occupancy fitted with Hill-Langmuir model. The solid black line shows the model fit, and bootstrap-derived 95% confidence intervals are shown in grey shading. Circles indicate peak scans; triangles show late scans connected to corresponding peak scans (dotted lines). Model fitting used peak scan data only. C, Receptor occupancy versus subjective drug intensity during PET scanning. The curve is fit with a sigmoid function. Each color represents an individual participant. Model fitting used baseline and peak scan data only (n=7 for peak scans).

Additionally, we assessed whether occupancy remained high after PLL had dropped using late-phase scans in two participants. P4 was administered 75 µg. At the peak scan (+161-281 minutes), they showed 84% occupancy and 8.5/10 SDI. At late scan (+420-540 minutes), they showed 74% occupancy, yet only 1.5/10 SDI, suggesting a temporal dissociation between receptor occupancy and subjective drug effects. P5 was administered 100 µg. At peak scan (+271-391 minutes), they showed 67% occupancy and 8.5/10 SDI. At late scan (+522-642 minutes), they showed 12% occupancy and 1/10 SDI, demonstrating that receptor occupancy eventually returns to near-baseline levels.

### LSD increases cerebral blood flow and internal carotid artery flow

Haemodynamic quantification was performed 30-60 minutes after the start of the PET scan. LSD intake produced widespread increases in CBF. Global CBF increased by 19.3% (Cohen’s dz = 1.20), with the strongest effects in the occipital cortex (26.4% increase; Cohen’s dz = 1.85) and thalamus (19.4% increase; Cohen’s dz = 1.04). We observed more modest CBF increases in limbic regions, including the hippocampus (15.1% increase; Cohen’s dz = 0.88) and amygdala (10.9% increase; Cohen’s dz = 0.36). Convergent with these CBF changes, ICA flow increased by 28% (Cohen’s dz = 1.30); yet ICA diameter showed negligible changes (-3.5%; Cohen’s dz = -0.38), suggesting the increased blood flow was not driven by macrovascular changes. The correspondence between CBF and ICA flow is shown in Supplementary Figure S5. Full details on regional responses are in Supplementary Table S2 and Figures S6 and S7. Single-subject CBF delta maps (changes in cerebral blood flow following LSD administration) are in Supplementary Figure S8.

### LSD decreases global brain connectivity

Each participant completed 1-3 10-minute multiband multi-echo fMRI-BOLD-fMRI scans during each PET scan. GCOR (i.e., the average correlation from any region to all other regions) was calculated for each scan and averaged within sessions to provide an estimate of brain connectivity. LSD decreased GCOR across most of the neocortex, with the largest decreases in visual and posterior default-mode network regions (Figure 3A, C, Supplementary Table S3 including results following global signal regression). In contrast, psilocybin increased GCOR across most regions with the strongest effects in the dorsal and ventral attention, control, and default-mode networks (Figure 3B, D). See Supplementary Figure S9 for single-subject GCOR change maps following LSD, including from late scans. Following LSD administration, we observed increased normalised spatial complexity (average dz = 0.56, range = 0.26 - 0.77 across networks), small-worldness (dz = 0.71), and sample entropy (average dz = 0.49, range = 0.03 - 0.87 across networks) and decreased geodesic entropy across a range of thresholds (dz range = -0.44 to -0.75). We also observed changes in network connectivity that mirror our GCOR findings and very small effects on signal complexity and modularity. For a complete overview of fMRI results with and without global signal regression, see Supplementary Table S4.

**Figure 3.**
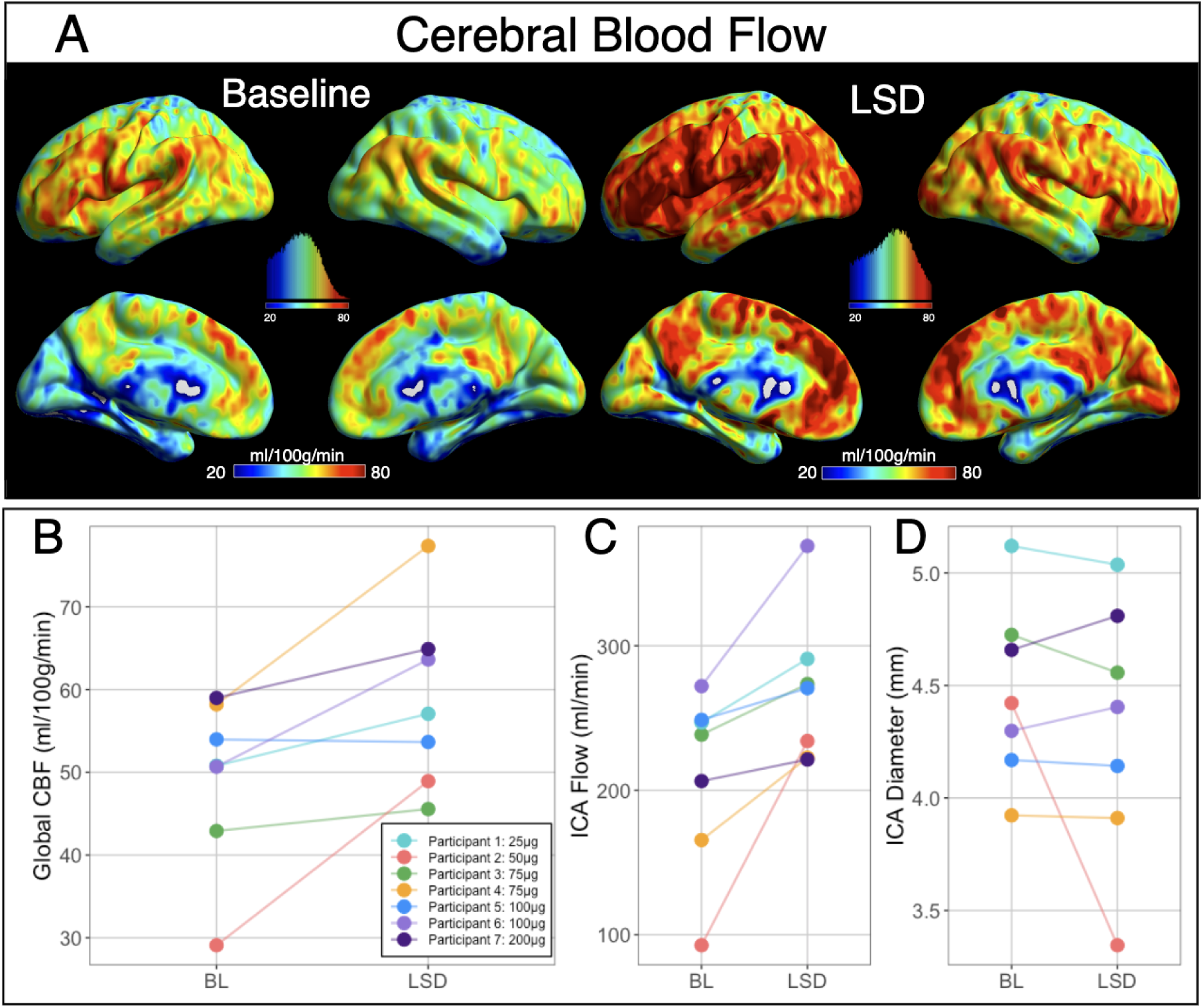
| Changes in cerebral blood flow (CBF) and haemodynamic parameters following LSD administration. A, Group-averaged CBF maps at baseline and after LSD administration, respectively. Color bars indicate CBF values in ml/100g/min. Histograms show distributions of voxel-wise values from the whole brain with the same scale as the colour bar below. B, Global CBF at baseline and post-LSD for individual participants. C, Internal carotid artery (ICA) flow at baseline and post-LSD. D, ICA diameter. In panels B-D, each color represents the same participant across all measurements, with lines connecting baseline (BL) and post-LSD (LSD) values.

### Correlations between global connectivity, change in cerebral blood flow, and receptor occupancy

To evaluate the relation between each neuroimaging metric, we calculated Spearman’s rank correlation coefficients (ρ) between the observed change in CBF, global connectivity across brain regions (GCOR), and neocortical 5-HT2AR occupancy across participants in each of 12 brain regions (Figure 5). GCOR was negatively correlated with neocortical occupancy across almost the entire neocortex, with 11 out of 12 regions showing negative correlations, of which seven were strong (ρ < -0.5) (Figure 5D). In contrast, change in CBF showed no clear relation with neocortical occupancy, with |ρ| < 0.2 in 10 out of 12 regions (Figure 5C). The relation between GCOR and CBF was predominantly negative, especially across large cortical regions. (Figure 5A). In the independent psilocybin cohort, we observe a similar negative correlation between GCOR and CBF, despite psilocybin producing an increase in GCOR (see Figure 4B) concurrent with decreased CBF (Figure 5B) (26) .

**Figure 4.**
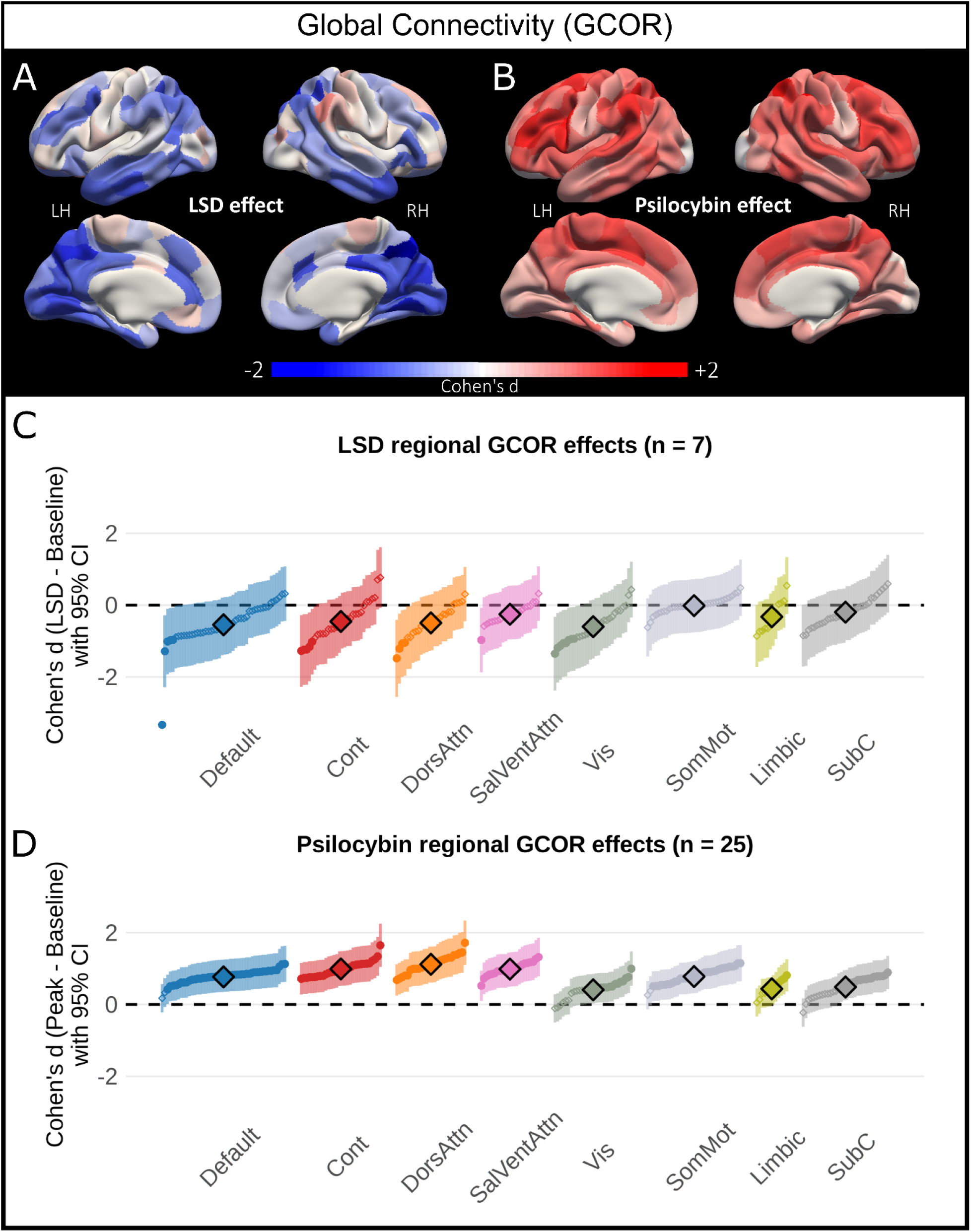
LSD and psilocybin produce opposing effects on global connectivity. A, Cortical surface maps showing LSD-induced decreases in global connectivity (GCOR) across widespread cortical regions (n = 7). B, Psilocybin-induced increases in GCOR showing a distinct spatial pattern predominantly affecting association cortices (n = 25). Colour scale represents Cohen’s d effect sizes ranging from -2 (blue, decreased connectivity) to +2 (red, increased connectivity). LH, left hemisphere; RH, right hemisphere. C, Forest plot displaying LSD effects on regional GCOR across functional networks. Points represent Cohen’s d effect sizes with 95% confidence intervals (shaded regions). Dashed line indicates no effect (d = 0). D, Forest plot displaying psilocybin effects on regional GCOR across the same functional networks, demonstrating increases across Default, Control, Dorsal Attention and Salience/Ventral Attention networks. Cont, Control network; DorsAttn, Dorsal Attention network; SalVentAttn, Salience/Ventral Attention network; Vis, Visual network; SomMot, Somatomotor network; Limbic, Limbic network; SubC, Subcortical regions.

**Figure 5.**
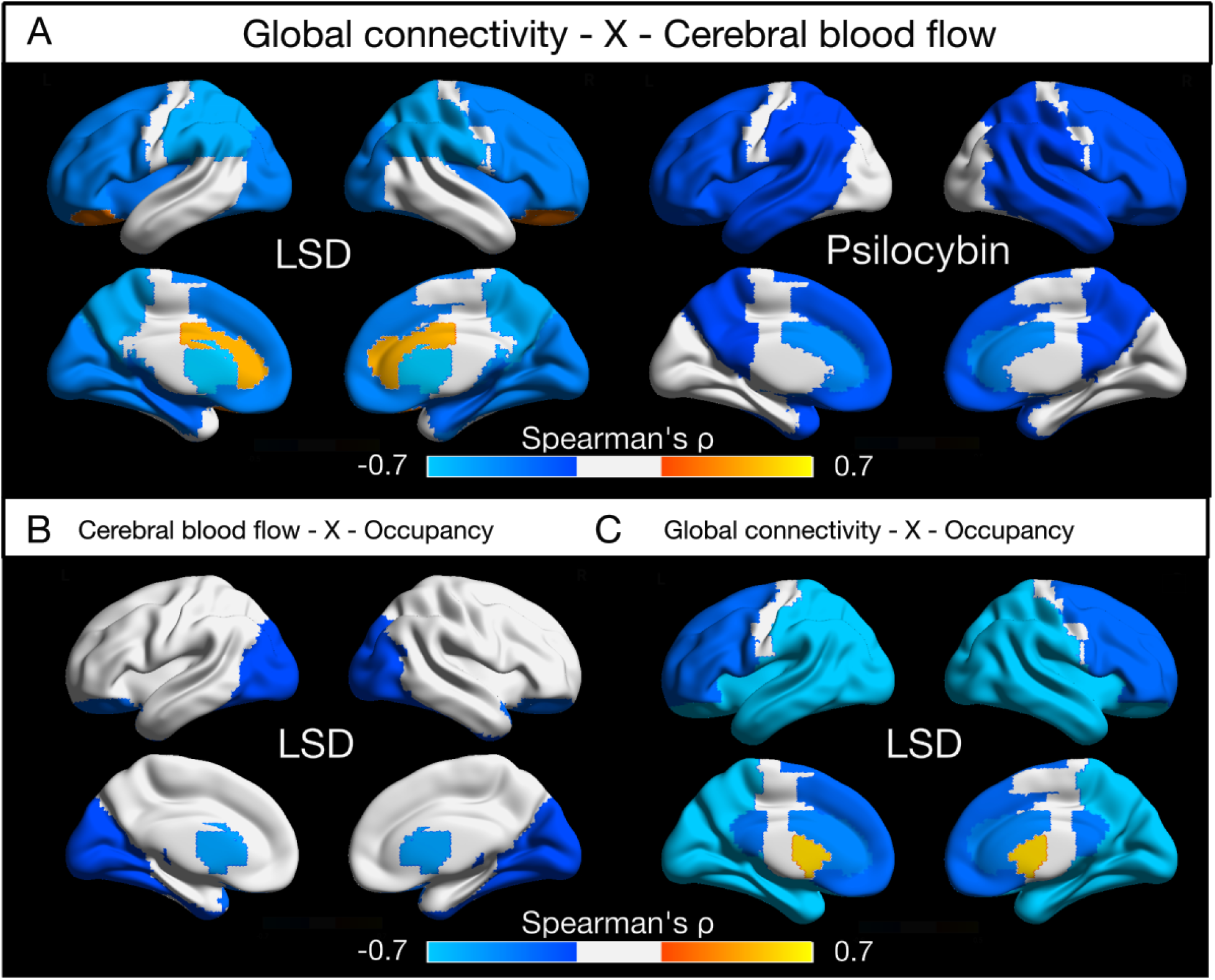
| Correlations between neocortical 5-HT2AR occupancy, cerebral blood flow, and global connectivity. Brain surface visualisations show Spearman’s rank correlations mapped to 12 AAL3 atlas brain regions: prefrontal, parietal, temporal, and occipital cortex, anterior cingulate cortex (ACC), posterior cingulate cortex (PCC), orbitofrontal cortex (OFC), putamen, insula, amygdala, caudate, and hippocampus. A, Regional correlations between global connectivity (GCOR) and cerebral blood flow (CBF) changes for LSD (left) and psilocybin (right). B, Regional correlations between CBF changes and neocortex 5-HT2A receptor occupancy for LSD. C, Regional correlations between GCOR changes and neocortex occupancy for LSD. Lateral (top) and medial (bottom) views shown for each comparison.

## Discussion

In this first human study to administer a psychedelic and acquire simultaneous molecular and functional brain imaging data, we characterised the pharmacodynamic effects of LSD in healthy volunteers, providing *in vivo* cerebral 5-HT2AR occupancy data and quantifying widespread alterations in CBF and functional brain activity. LSD exhibited high-affinity binding to cerebral 5-HT2AR and hysteresis effects between plasma drug levels and subjective drug effects, increased global CBF substantially, and increased flow in the internal carotid arteries without inducing changes in the diameter. LSD decreased GCOR, this change was negatively associated with change in CBF. We contrast our findings using an independent dataset of participants administered psilocybin. Psilocybin does not produce a hysteresis effect between plasma drug levels and subjective drug intensity and increases GCOR. As is the case for LSD, for psilocybin the change in GCOR was similarly negatively associated with change in CBF.

### Comparison with *in vitro* work and future applications

Our receptor occupancy results, with an IC_50_ of 1.9 nM, align well with prior *ex vivo* receptor binding studies conducted on human cortical tissue: specifically, previous work using the antagonist radioligand 3H-spiperone reported an affinity of 4 nM (27) and 3H-ketanserin in 5-HT2AR-expressing NIH-3T3 cells indicated an affinity of 4.2 nM (28). For comparison, clinical data suggest that the EC_50_ for psychedelic effects is 3-4 nM (29). This convergence across distinct methodologies is reassuring and further validates [^11^C]Cimbi-36 PET as a tool for evaluating target engagement in humans *in vivo*, relevant for characterising other forthcoming 5-HT2AR-based therapeutics. This is especially important for the Phase 1 testing of prospective non-psychedelic 5-HT2AR agonists, for which target engagement cannot be estimated using behavioural measures. Consistent with our observations assessing psilocybin’s 5-HT2AR occupancy (30), we observed a generally positive relation between 5-HT2AR occupancy and self-reported subjective drug intensity. However, we observed notable inter- and intra-participant variation across estimates, with one participant receiving only 25 μg LSD rating 10/10 subjective intensity with a corresponding occupancy of 42%, and another rating only 1.5/10 SDI despite having 72% receptor occupancy during a late scan. This positive, yet variable, relation indicates that clear evidence of target engagement requires molecular imaging; it cannot necessarily be derived from subjective effects.

As only the second psychedelic compound after psilocin (30) to have its receptor occupancy profile characterised in humans, this work establishes a framework for evaluating target engagement of emerging compounds. Several classical psychedelics, including 5-MeO-DMT (NCT05800860, NCT05839509) and N,N-DMT (NCT06094907, NCT05553691), are advancing in clinical development without direct occupancy measurements. MDMA has progressed through phase 3 trials (31,32), and some of its clinically relevant effects are known to be blocked by the 5-HT2AR antagonist ketanserin (33). Yet, the degree to which the R-enantiomer of MDMA occupies the 5-HT2AR either directly or via R- and S-enantiomer mediated endogenous serotonin release is unresolved, could be resolved using PET, and may be critically relevant for understanding its efficacy and safety.

### The relation between dose and occupancy

Our model provides a valuable tool for translating measured plasma LSD concentrations into estimated 5-HT2AR occupancy levels. Mechanistic work based on plasma levels alone overlooks the non-linear relation between drug concentrations and receptor binding. By enabling the conversion from plasma measurements to receptor occupancy estimates, our model offers a more mechanistically sound approach for interpreting experimental and clinical outcomes in LSD research, potentially improving correlations with biological and clinical endpoints, including physiological biomarkers, subjective experiences, therapeutic outcomes, and neuroimaging measures. Based on typical peak plasma concentrations (4), oral doses of 25 µg, 50 µg, 100 µg, and 200 µg yield approximate receptor occupancies of 43%, 62%, 74%, and 84%, respectively. However, we and others have observed substantial inter-individual variation in peak plasma levels, insufficiently explained by bodyweight or *CYP2D6* genotype (34); occupancy cannot be reliably estimated from dose alone but requires at least blood samples to be measured for drug concentration over time. Our model relating bodyweight-adjusted dose to 5-HT2AR occupancy indicates that a dose of 1 µg/kg LSD (freebase) will produce ∼70% 5-HT2AR occupancy, which is sufficient for maximal subjective drug effects while limiting potential off-target effects.

Our PET data shows an Occ_max_ of 97% for LSD - notably higher than previously reported values for other 5-HT2AR ligands including psilocin (77%) (30), ketanserin (77%) (35), volinanserin (77%) (36), and pimavanserin (78%) (37). Future work administering high doses of LSD and psilocybin may elucidate whether this observed difference in maximal occupancy is related to consequent differences in downstream behavioural or neural effects. However, the confidence of our Occ_max_ estimate is constrained by limited sampling at high plasma concentrations. The increasing prevalence of adverse effects such as nausea and anxiety at higher doses may pose inherent challenges for future high-dose occupancy studies. Future studies may also wish to account for the concentration of the LSD metabolite nor-LSD, which has been shown to be an agonist at 5-HT2AR with a K_i_ of approximately 6.1 nM (25).

All but one participant showed a clear anticlockwise hysteresis loop between plasma LSD levels and subjective drug effects, such that SDI peaked approximately half an hour after PLL, conversely, we did not observe this effect with psilocybin. Such a delay could be due to slow brain penetrance, post-receptor signalling, or receptor binding. However, LSD has biochemical properties that make slow brain penetrance unlikely (38) and has similar post-receptor signalling properties to psilocybin (39). Structural studies have demonstrated that LSD exhibits unusually slow dissociation kinetics from 5-HT2AR due to unique hydrogen-bond interactions, the so-called “lid-hypothesis”, which may result in the gradual accumulation of 5-HT2AR-bound LSD, thus explaining the observed delay (14). This slow dissociation is not observed with psychedelics from other structural classes (15). This hypothesis could be tested using either radiolabeled LSD or carefully timed [^11^C]Cimbi-36 scans in the early phase of drug effects. The temporal continuity of our SDI ratings may also be biased by the fact that participants changed from the quiet treatment room to the scanner environment around peak drug effects; however, most participants were already at peak subjective drug effects when transported to the scanner.

We observed one participant who, despite a significant reduction in subjective drug effects at their late-scan, retained high occupancy of 5-HT2AR (in line with their PLL), implying rapid post-receptor tolerance effects, which may be evaluated preclinically. PET cannot distinguish between reductions in available receptor binding sites due to occupancy by cold ligand (i.e, LSD), or by receptor downregulation; therefore, it may be that the observed occupancy is the result of receptor downregulation. However, previous PET work with psilocybin showed no reduction in 5-HT2AR binding one week after a high dose of psilocybin (40), and we show in one participant that BP_ND_ returns almost to baseline after only 8 hours, indicating no substantial rapid receptor downregulation that would positively bias our occupancy estimates. Psilocybin similarly showed a return to baseline BP_ND_ once the drug was cleared (30). We are limited to only qualitatively presenting data from the late scans due to losing three scans because of scanner failures and adverse effects, limiting our capacity to test the lid-hypothesis as described in the preregistration.

### Haemodynamic effects of LSD vs psilocybin

Our observation of increased global CBF (19.3%) under LSD stands in marked contrast to the cerebral haemodynamic effects of psilocybin, for which four studies have reported reduced CBF, in absolute units (ml/100g/min) (26,41–43). One study found global CBF reductions of 12% following psilocybin administration, with regional analysis revealing the strongest decreases in parietal (-15%) and occipital regions (-14%), alongside significant ICA constriction (-7%) (26). Following LSD, we observed widespread CBF *increases*, most pronounced in the occipital (26%) and parietal cortices (18%). These CBF changes were accompanied by increased ICA flow (28%), with ICA diameter remaining relatively stable. The only previous neuroimaging study of LSD’s effects on CBF employed 2D time-series acquisition following intravenous LSD administration and reported elevated occipital blood flow. However, the absolute CBF values reported in that study exceeded physiological ranges (>150 ml/100g/min) in both the LSD and placebo conditions, limiting their interpretability (13,23).

LSD and psilocybin both produce their psychedelic effects primarily via 5-HT2AR agonism, yet these opposing effects on cerebral haemodynamics suggest distinct downstream or off-target mechanisms. This is supported by our observation that, unlike GCOR effects, 5-HT2AR occupancy was not correlated with changes in CBF. LSD binds with nanomolar affinity to several monoaminergic receptors, including most serotonin, adrenergic alpha 2A and dopamine subtype 2 receptors, whereas psilocybin’s receptor binding profile is mostly restricted to the serotonin receptors (28,44). Both psilocybin and LSD are agonists at 5-HT2AR and 5-HT1BR, which induce vasoconstriction (45,46); yet, LSD does not produce vasoconstriction at physiological levels *in vitro,* aligned with our observations here (47). LSD therefore may activate counter-acting vasodilatory pathways, which can be evaluated in future research with selective antagonists. The observed increase in CBF is unlikely to be driven by systemic confounds; although LSD produces increases in blood pressure (4), these are well within the range that is accounted for by autoregulation (48). A [^18^F]FDG-PET study has previously shown that psilocybin increases cerebral glucose metabolism despite concurrent reductions in blood flow, implying a functional decoupling between neural activity and perfusion, which may be due to 5-HT2AR-mediated vasoconstriction, as discussed above (49). This hypothesis may be evaluated using a peripherally selective 5-HT2AR antagonist such as sarpogrelate (50). Whether such decoupling also applies to LSD remains unknown, as [^18^F]FDG-PET imaging has not yet been performed. Simultaneous [^18^F]FDG-PET/MR could directly evaluate this relation between metabolism, blood flow, and neural activity following LSD administration. Previous work with psilocybin has also not yet evaluated ICA flow, which can be measured on all major MRI systems in under two minutes using a phase-contrast mapping sequence.

### Effects of LSD on functional brain connectivity

Three previous studies have evaluated LSD effects on GCOR. One reported increases across fronto-parietal, default-mode and salience networks (n = 15) (51), another primarily decreases across the visual and somatomotor regions (i.e., without global signal regression, as evaluated here) (n = 24) (10), and one reported decreases in the visual cortex and increases in striatal regions (n = 28) (52). Our observations are consistent with decreases in visual cortex GCOR as well as across dorsal attention, control and default-mode network regions. These decreases in GCOR were strongly correlated with neocortical 5-HT2AR receptor occupancy and are thus likely an on-target effect. We contrast our findings with an independent cohort of participants administered psilocybin, showing increased GCOR across most of the cortex. Because of the experimental design employing varying doses of LSD and a small sample size, which were chosen based on the occupancy outcomes, we are constrained in our capacity to infer statistically significant effects on functional neural effects. These fMRI findings nevertheless contribute valuable new data to the field, particularly given that most previous analyses of LSD effects have evaluated the same dataset and extremely few studies have evaluated previously investigated methods (18). Presentation of strictly thresholded statistical maps can exaggerate inconsistencies across studies (53); therefore, we report effect sizes from all analyses, which can help establish which neural signatures of LSD are most deserving of continued investigation and facilitate future meta-analyses and cross-study comparisons (54).

We observed a negative correlation between LSD-induced increases in CBF and decreases in GCOR. We observed the same negative relation in our psilocybin cohort that showed increases in GCOR and decreases in CBF. The neurovascular coupling model relates increased cerebral metabolic activity with blood flow overcompensation and subsequent increases in the BOLD signal (55,56). By altering CBF, psychedelics may modulate this coupling, as has been reported for, e.g., caffeine (57). A preclinical evaluation of the effects of the psychedelic DOI has shown the potential for 5-HT2AR mediated alterations in neurovascular coupling (58). Hypercapnia increases CBF without affecting neural activity (59) and has been shown to reduce BOLD correlations, possibly by decreasing signal-to-noise (60), which would also theoretically increase measures of signal complexity, i.e., entropy. Our observations highlight the urgent need for a future study to characterise psychedelic effects on neurovascular coupling by relating cerebral metabolic, blood flow, and BOLD effects. If psychedelics are shown to affect neurovascular coupling, it would substantially affect the interpretation of psychedelic BOLD effects presented in dozens of previous studies, including those presented here.

In summary, this study provides the first *in vivo* pharmacodynamic profile of LSD in humans through simultaneous molecular and functional neuroimaging. We characterise LSD’s high-affinity binding to 5-HT2AR and present data suggesting a temporal disconnect between LSD plasma concentrations, receptor occupancy, and subjective drug effects that was not observed for psilocybin. Our findings of increased cerebral blood flow and decreased GCOR contrast markedly with psilocybin’s effects, suggesting pharmacological distinctions between these related psychedelics. We provide preliminary evidence that such alterations in connectivity may be confounded by hemodynamic effects, and propose future studies, especially involving direct measurement of the effects of LSD on cerebral metabolism, to help resolve the basis of the observed neural effects. The observations presented here advance our ability to dose LSD in clinical populations and progress our understanding of the neural mechanisms that may underpin their acute and lasting effects.

## Methods

### Ethics and informed consent

All research procedures were conducted in accordance with the Declaration of Helsinki (61). The study protocol was approved by the ethics committee of the Capital Region of Denmark(H-21060056) and the Danish Medicines Agency (EudraCT no.: 2021-002633-42; CTIS: 2024-519564-41-00). Participants were given information packs including an overview of their rights and the use of their data following telephone screening and before signing the consent forms. The study was preregistered at https://clinicaltrials.gov (NCT05953038) and https://aspredicted.com (https://aspredicted.org/gn3un.pdf).

### Inclusion and exclusion criteria

Inclusion criteria were:

- Healthy individuals 18-75 years old

Exclusion criteria were:

- Current or past history of primary psychiatric illness (DSM-IV axis-I or WHO ICD-10 diagnostic classification)
- Current or past history of primary psychiatric illness (DSM-IV axis-I or WHO ICD-10 diagnostic classification) in a first-degree relative (i.e., parents, siblings)
- Current or past history of neurological disease, significant somatic condition/disease
- Use of medication that could potentially influence results (e.g., drugs that act on relevant components of the serotonin system or may interfere with the metabolism of LSD)
- Non-fluent Danish language skills
- Profound visual or auditory impairments
- Severe learning disability
- Pregnancy on the scan date, verified by a pregnancy test (females, test omitted if confirmed that individual is post-menopausal)
- Breastfeeding (females)
- Contraindications for MRI (e.g., pacemaker, claustrophobia, etc.)
- Contraindications for PET (e.g., claustrophobia, radiation exposure, etc.)
- Current alcohol or drug abuse
- Allergy to administered compounds
- Participant in research study with >10 mSv exposure within the past year or significant occupational exposure to radioactive substances
- Abnormal ECG (i.e., indicating current, previous heart disease or predisposition thereof, e.g., QT prolongation) or use of QT prolonging medication
- Use of psychedelic substances within the preceding six months
- Blood donation up to three months before the study (i.e., more than 500 ml of blood)
- Head injury or concussion resulting in loss of consciousness for more than 2 minutes
- Haemoglobin levels < 7.8 mmol/l for women and 8.4 mmol/l for men
- Ferritin levels outside normal range (12-300 μg/L)
- Body-weight < 50 kg or > 110 kg
- BMI > 35 kg/m²
- Individual assessment by research staff deeming drug administration unsafe due to ethical or psychological circumstances of the participant

### Recruitment

Healthy volunteers were recruited from an in-house database of individuals interested in participating in a study involving psychedelics (see CONSORT diagram Supplementary Figure S10). A total of 52 volunteers were contacted via telephone, 32 were excluded at telephone pre-screening, two declined before in-person screening, three were excluded during in-person screening, and 2 dropped out after inclusion but before any study visits. The data presented here are from 11 healthy participants collected between November 2023 and November 2024. Details of recruitment, procedures during the LSD session, ethical approvals, MRI acquisition, and quality control are described in the Supplementary Text. Analyses were pre-registered on the 27th of February 2023 (https://aspredicted.org/gn3un.pdf). No statistical methods were used to pre-determine sample sizes.

### Study Design

See Supplementary Figure S11 for an overview of scan elements.

Visit 0: Following telephone screening and informed consent, participants underwent a screening battery including evaluation of medical and drug-use history, pregnancy, electrocardiogram, neurological exam, and psychiatric health.

Following inclusion, participants were asked to fill out, online from home, a 2-hour set of state and trait questionnaires.

Visit 1: Whole-blood, plasma, and buffy-coat samples were taken, as well as a urine sample to check for the absence of drugs of abuse and for women to check for pregnancy. Participants then filled out a collection of state questionnaires and underwent a 2-hour simultaneous PET and MRI scan. Music was played via earphones during the entire scan, except when questions were asked. Scan parameters are discussed below.

Visit 2 involved a preparatory conversation with the psychological support staff, typically lasting 60-90 minutes, focused on the LSD intervention process. The conversation addressed both the psychological and physiological effects of the intervention, the role of the support staff, and the participants’ expectations for the session. All participants were informed about changes they might experience in perception, emotions, and sense of self, and were provided with techniques to navigate these shifts with openness and acceptance, as well as guidance on managing possible physical effects such as nausea or dizziness. The conversation also explored participants’ past experiences with psychoactive substances, and practical matters, including transportation, accommodation arrangements, and post-session support, were addressed to ensure participants had a reliable support network in place. Participants were reminded of the rules and safety measures, including consent for touch and the prohibition of certain behaviours. Additionally, agreements regarding communication during the scanning process were discussed to ensure participants understood when and how to communicate with the support team. Occasionally, this conversation occurred before the baseline PET scan. Three participants required an additional preparatory session. One was needed because the initial conversation had taken place more than two weeks before the intervention. Another participant required extra preparation to familiarise themselves with the support team, and one participant needed an extra session to integrate talking points from the first preparatory conversation.

Visit 3: 1 week to 1 month later, participants came in for their intervention session. Participants were fitted with two venous catheters, gave blood samples, filled out the same state questionnaires as during baseline scans, and received a dose of LSD tartrate between 25 μg and 200 μg equivalent as LSD freebase. Participants, but not study staff, were blinded to the dose received. Participants were dosed in a comfortable aesthetic room with speakers and in the presence of two trained psychological support staff. Approximately 1 h 45 m following drug administration, participants were transported via wheelchair to the PET-MR scanner. They then underwent the same scan as at visit 1, with injection of radiotracer at approximately 2 h 45 m after LSD administration, such that tracer injection was approximately aligned with peak subjective drug effects. Two participants (P4 and P5) received late scans at 7 h 10 mins and 8 h 42 mins respectively. Following the scan, participants returned to the comfortable room via wheelchair. They remained there until their subjective drug effects had diminished. Then they drew a mandala of their experience and were either picked up by a friend or family member and returned home, or were escorted by study staff to the patient hotel at the hospital.

Visit 4: Either the next day or the day after, participants returned for a follow-up conversation with the psychological support staff. From home, participants filled out an online deck of questionnaires describing their acute drug effects. One participant had an extra integrative phone conversation with a member of the psychological support staff.

Three months after intervention, participants were sent a similar online deck of trait and state questionnaires as they filled out at visit 0, which were filled out at home. Participants were instructed to report all adverse effects to the study staff irrespective of whether they deemed them related to the study medication. All staff with participant contact were responsible for the identification of potential adverse effects, which were eventually evaluated by a medical doctor.

### PET scanning parameters

PET images were acquired for 120 minutes on one of two identical 3T Siemens Biograph mMR scanners (Siemens Healthcare, Erlangen, Germany) after a bolus injection of [^11^C]Cimbi-36. Processing of [^11^C]Cimbi-36 PET data was performed as described by (30). PET data was reconstructed using MR-imaging based attenuation-correction (DeepDixon) (62) and a framing protocol of 6x10s, 6x20s, 6x60s, 8x120s, 19x300s. All frames were aligned to frame 27, with frames 1-12 using the transformation parameters calculated from frame 13’s alignment. Segmentation was performed using SPM12MultiSpectral segmentation using session-specific high-resolution T1- and T2-weighted MRI scans. Regions of interest were defined using PVElab (63), a fully automated regional delineation procedure, and regional time-activity curves were extracted for kinetic modeling.

Kinetic modeling was performed using the simplified reference tissue model (SRTM) with neocortex (a volume-weighted average of the below cortical regions) chosen a priori as the region of interest due to the high expression of 5-HT2ARs and the beneficial signal-to-noise ratio within this region; cerebellum was chosen as the reference region due to its absence of 5-HT2AR in humans (64).

- Orbital frontal cortex
- Medial inferior frontal gyrus
- Superior temporal gyrus
- Parietal cortex
- Medial inferior temporal gyrus
- Superior frontal gyrus
- Occipital cortex
- Sensorimotor cortex

Nondisplaceable binding potential (BP_ND_) (65) was the primary outcome measure, with occupancy % calculated as 100 x ((baseline BP_ND_ - LSD scan BP_ND_)/baseline BP_ND_). The relation between plasma LSD concentration and receptor occupancy was modeled using the single-site binding model, the Hill-Langmuir equation:

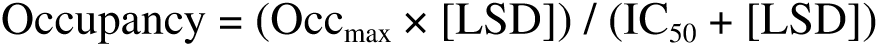

where Occ_max_ represents maximum achievable occupancy and IC_50_ represents the plasma concentration producing 50% occupancy, and [LSD] was the average concentration of LSD from blood samples taken during the PET scan (one at injection, two during the scan, and one at scan cessation).

The Hill-Langmuir equation parameters were estimated using nonlinear least squares regression via the stats::*nls* function (version 4.3.1) in R with the ‘port’ algorithm, which ensures physically meaningful parameter estimates through constrained optimisation (i.e., Occ_max_ bounded between 0-100%, IC_50_ constrained to positive values). To assess uncertainty in these estimates, we performed a residual bootstrap with 10,000 iterations. For each iteration, residuals from the observed model were randomly resampled with sign flips and added to the fitted values to generate new synthetic datasets preserving the underlying relation. The model was fit to each synthetic dataset using the same constrained optimisation approach. This procedure provides robust confidence intervals that respect the physical constraints of receptor binding while accounting for the uncertainty in our limited sample size. Bootstrap estimates were calculated using the median of the bootstrap distribution, with 95% confidence intervals defined by the 2.5th and 97.5th percentiles.

Surface maps of 5-HT2AR BP_ND_ (Figure 2) were generated in PETSurfer using MRTM2, where k2’ was estimated with neocortex as the high-binding region and cerebellum as the reference region (66). These maps are used only for data visualisation. All other reported BP_ND_ or occupancy estimates are from SRTM-derived BP_ND_ values.

### MRI scanning parameters

During the entire baseline and first post-intervention PET-MRI scan, participants listened to the following playlist: https://open.spotify.com/playlist/5yT9nTRdgoPIb2f0h5QuKg?si=b6374a44413a485b; participants who completed a second PET-MRI scan after LSD administration listened to the following playlist: https://open.spotify.com/playlist/0UW0wjkE14sXIjwqlmKf5Y?si=74ad7745384244c5.

During the 120-minute PET scan, we acquired the following MR scans using a 32-channel head coil:

High-resolution T1-weighted structural images with a 3D magnetization-prepared rapid gradient-echo (MPRAGE) sequence with the following parameters: repetition time (TR) = 2300 ms, echo time (TE) = 2.26 ms, inversion time (TI) = 900 ms, flip angle = 8°, field of view (FOV) = 256 x 256 mm^2^, matrix size = 256 x 256, voxel size = 1 x 1 x 1 mm^3^, 176 sagittal slices, and GRAPPA parallel imaging with an acceleration factor of 3.

High-resolution T2-weighted structural images were acquired using a 3D turbo spin-echo sequence (SPACE) sequence with the following parameters: repetition time (TR) = 3200 ms, echo time (TE) = 407 ms, variable flip angle with a nominal value of 120°, field of view = 230 x 230 mm^2^, matrix size = 256 x 256, 0.9 mm isotropic voxels, 256 sagittal slices, GRAPPA in-plane acceleration factor = 2, partial Fourier sampling, echo-train length = 254, receiver bandwidth = 725 Hz/pixel; images were reconstructed to a 512 x 512 matrix.

We acquired field maps using the vendor-provided gradient echo field mapping sequence with the following parameters: TR = 500 ms; TE1/TE2 = 6.16/8.62 ms; flip angle = 60°; FOV = 100% phase FOV; matrix size = 68 × 68; slice thickness = 3 mm with 3 mm spacing; phase encoding direction = anterior-posterior (j-). The sequence produced three images: magnitude images at both echo times and a phase difference map calculated between the two echoes.

These were then followed by either two or three functional imaging blocks, time permitting, consisting of the following scans:

Pseudo-continuous arterial spin labelling (pcASL) images were acquired using a five post-label delay 3D turbo gradient spin echo sequence with the following parameters: scan time = 7 min 11 sec, label/control pairs = 12, native voxel size = 3 x 3 x 3 mm, image matrix = 96 × 96, number of slices = 40, label duration = 1508 ms, post-label delays = [500, 500, 1000, 1000, 1500, 1500, 2000, 2000, 2000, 2500, 2500, 2500] ms, echo time = 3.78 ms, repetition time = 4100 ms, flip angle = 120°), two background suppression pulses optimised for each PLD. The acquisition included an M0 scan with which to calibrate the pcASL signal without any background suppression.

Time of Flight (TOF) angiography images were acquired using a 3D gradient recalled (GR) sequence with the following parameters: scan time = 6 min 30 s, slice thickness = 0.8 mm, echo time = 4.79 ms, repetition time = 27 ms, flip angle = 18°, acquisition matrix = 482x640 covering a 150x200 mm^2^ field-of-view.

Phase Contrast Mapping (PCM) images were acquired using a 2D sequence with the following parameters: scan time = 8.36 s, slice thickness = 8 mm, echo time = 7.5 ms, repetition time = 100.4 ms, flip angle = 10°, VENC = 100 cm/s, acquisition matrix = 320x320 matrix covering a 240x240 mm^2^ field-of-view.

BOLD functional images were acquired using a multi-echo, multi-band echo-planar imaging (EPI) sequence with the following parameters: TR = 1117 ms, number of volumes = 550 (10 mins 14 s), TEs = 15.0/33.68/52.36 ms, flip angle = 70°, FOV = 204 x 204 mm, matrix size = 68 x 68, voxel size = 3 x 3 x 3 mm, axial slices = 42, multi-band acceleration factor = 3, in-plane acceleration (GRAPPA) acceleration factor = 2, and phase-encoding direction = anterior-to-posterior (j-).

### Investigational medicinal product

The study medication was produced by Apotheke Dr. Hysek, Biel, Switzerland, in accordance with Good Manufacturing Practice (GMP). LSD was prepared as drinking solutions containing 36.5 µg or 146 µg of GMP-grade LSD tartrate (Lipomed AG, Arlesheim, Switzerland), corresponding to 25 or 100 µg of LSD base, dissolved in 1 mL of 20% alcohol solution (v/v). Participants were blinded to dose by affixing opaque stickers to the bottles before being given to the participants. Participants were not blinded to the fact that they would receive LSD. The investigational medicinal product was stored in a locked safe within a fridge kept at 4-6℃, and doses were prepared on the actual dosing day.

### Behavioural effects

Before each EPI scan, participants were asked to listen to the music. Immediately following each scan, the participants were first asked if they had fallen asleep, and then to rate the following questions on a scale from 0 to 10, where 0 was *not at all* and 10 was *very much*: How calm do you feel? How much claustrophobia are you experiencing? How tired do you feel? How much physical discomfort are you in? Participants were then asked to evaluate the effect of the music they listened to during that scan on the following emotions from the Geneva Emotional Music Scale (GEMS): wonder, transcendence, tenderness, nostalgia, powerfulness, happiness, peacefulness, sadness, and tension.

On the dosing day, participants were asked to rate how intense their LSD experience was on a scale from 0 to 10 approximately every 20 minutes from ingestion of the drug until they were considered sober by study staff. During scans, these were less frequent, aligning with blood samples.

### Cerebral blood flow quantification

The preprocessing pipeline of pcASL consisted of several steps. First, label/control difference images were realigned for each post-label delay (PLD). The high-resolution T1-weighted structural image was then co-registered with the M0 image. Absolute CBF quantification was performed using BASIL (part of FSL - FMRIB software library) on all pcASL images (67). This process involved label/control image subtraction and signal calibration to absolute CBF (ml/100g/min), incorporating M0 scan data and employing a single, well-mixed tissue compartment model for tracer kinetics modeling. Standard parameters were applied for the quantitative modeling process (23). The structural image underwent segmentation using SPM12 to generate probability maps for grey matter, white matter, and cerebrospinal fluid. Subsequently, the calibrated pcASL image was transformed into MNI standard space using warps estimated during the segmentation step. Finally, the normalized pcASL images were smoothed spatially using a 3D Gaussian kernel (8 mm full-width half-maximum (FWHM)).

For regional analyses, we used the Anatomical Automatic Labelling atlas version 3 (AAL3) (68), which provides detailed cortical and subcortical parcellation in MNI space. We combined anatomically adjacent regions to create larger, functionally relevant regions of interest (ROIs). This resulted in 13 bilateral regions: prefrontal cortex, temporal cortex, parietal cortex, occipital cortex, anterior and posterior cingulate cortices, thalamus, amygdala, putamen, caudate nucleus, hippocampus, insula, and orbitofrontal cortex. Regional CBF values were calculated using grey-matter weighted averaging based on subject-specific segmentation maps. The specific composition of these compound ROIs followed the same approach as previously described (26), with full details provided in the Supplemental Table S5. We focused our analyses on absolute CBF values, retaining the quantitative units (ml/100 g/min). In our within-subject, repeated-measures design, each participant serves as their own baseline, allowing us to estimate regional drug-related changes in absolute CBF.

### Internal carotid artery diameter quantification

We quantified ICA diameter using publicly available in-house software (https://github.com/kristian1801/ica-segmentation-tool). Through this software’s pipeline, the TOF images were automatically processed with background subtraction, anisotropic diffusion filtering, and top-hat filtering to enhance vascular structures. The software’s graphical interface allowed manual selection of the ICA, which was then automatically segmented using a 2D region-growing algorithm with dynamic intensity thresholding across axial slices. The software then reconstructed a 3D model of the segmented ICA and calculated vessel diameter using the median of measurements from each slice. All images were visually inspected to ensure the quality of the estimates.

### Internal carotid artery flow quantification

We quantified ICA flow using previously developed PCMCalculator software (69,70) (software available at https://github.com/MarkVestergaard/PCMCalculator). Through this software’s pipeline, PCM images were processed to measure through-plane flow. The software’s graphical interface allowed manual selection of the ICA, which was then automatically analysed to calculate the flow rate. The methodology has been previously validated for both 3T Philips and 3T Siemens MR systems.

### pcASL data quality control

pcASL and fMRI quality control are graphically described in Supplementary Figure S12.

#### 1. Initial Data Collection and Exclusions

The dataset included multiple pcASL runs from baseline and LSD intervention sessions Early quality assessment identified systematic signal dropout issues in runs 2 and 3, and 11 runs from 6 participants were excluded due to shimming-related signal dropouts.

#### 2. Registration Quality Control

All pcASL images underwent visual inspection before and after transformation to MNI space. Registration quality was assessed by:

- Alignment with participant structural images
- Correspondence with standard atlas landmarks
- Verification of anatomical boundaries and key structures.

#### 3. Final Dataset Construction

7 participants met all quality criteria for both baseline and LSD sessions Baseline CBF maps were constructed by averaging:

* 3 runs for 3 participants
* 2 runs for 4 participants

– LSD session analysis used only the first run from each participant’s session.

### Functional MRI preprocessing and denoising

Functional and anatomical preprocessing was performed using Configurable Pipeline for the Analysis of Connectomes (C-PAC version 1.8.7. Dev1). The .yml file containing all settings used is available on the project GitHub (https://github.com/Pneumaethylamine/dOccLS). T1-weighted anatomical images underwent brain extraction using FSL’s Brain Extraction Tool with the “robust” setting. Tissue segmentation using FSL’s FAST and segmentation masks were made using a probability threshold of 0.9. Functional preprocessing steps included slice-timing correction, motion correction using AFNI’s 3dvolreg, and distortion correction using a phase difference field map. Functional images were masked using AFNI and intensity-normalised. For spatial normalisation, anatomical images were registered to the MNI152 template using Advanced Normalization Tools (ANTs). Functional images were co-registered to the anatomical image using FSL’s FLIRT with boundary-based registration, and subsequently transformed to MNI space. Nuisance regression was performed in each subject’s native space. We applied ICA-AROMA (non-aggressive). Additional nuisance regressors included 24 motion parameters (6 motion parameters, their derivatives, squared terms, and squared derivatives), the first 5 principal components from white matter and cerebrospinal fluid, mean CSF signal, and linear and quadratic trends. A bandpass filter (0.01-0.1 Hz) was applied. Global-signal regression was not applied.

Preprocessed data were visually inspected at each step; following visual inspection, it was deemed that brain extraction was superior for one participant using the non-robust FSL BET setting, all other preprocessing steps remained the same. Post-processing steps included spatial smoothing using a 4mm FWHM Gaussian kernel, normalisation, and demeaning.

### Functional MRI data quality control

See Supplementary Figure S12 for an overview of quality control steps.

1. Initial Data Exclusions

– Early quality assessment revealed systematic signal dropout issues in runs 2 and 3 of several sessions related to erroneous shim settings
– 4 runs from 3 participants were excluded due to these shimming-related signal dropouts.
2. Preprocessing Quality Control

– Standard preprocessing was applied to all remaining runs
– During ICA-AROMA denoising, one run was classified as containing >90% noise components
– Visual inspection of this run confirmed severe artifacts throughout the timeseries
– This run was excluded from further analysis.
3. Motion Assessment

– Mean framewise displacement (FD) was calculated using Power’s method
– A threshold of 0.2 mm mean FD was set as the initial screening criterion
– All runs exceeding this threshold underwent comprehensive visual inspection, including:

* Temporal plots of 6 motion parameters and their derivatives
* DVARS timeseries
* Functional time series for key regions of interest
* Seed-based connectivity matrices
* Raw EPI data and preprocessed BOLD timeseries
– Based on this assessment, 2 runs showing excessive motion-related artifacts were excluded.
4. Final Dataset Construction

– 7 participants met all quality criteria for both baseline and LSD sessions
– Baseline scores were calculated by averaging:

3 runs for 4 participants
2 runs for 3 participants
– LSD scores were calculated by averaging

3 runs for 3 participants
2 runs for 2 participants
1 run for 2 participants

### Psilocybin data

Psilocybin data were drawn from a previously reported dataset (26,71). Please see the original publication for a description of data collection, preprocessing, and denoising beyond that described below.

#### Participants and study design

Twenty-eight healthy volunteers (10 female; mean age ± SD: 33 ± 8 years) participated in a single-blind, crossover study approved by the ethics committee of the capital region of Copenhagen (H-16026898) and the Danish Medicines Agency (EudraCT: 2016-004000-61). After providing written informed consent and medical screening, participants received a single oral dose of psilocybin (0.2–0.3 mg/kg; mean ± SD: 19.7 ± 3.6 mg). Resting-state fMRI scans were acquired before drug administration (predrug) and approximately 130 minutes post-administration (peak). Plasma psilocin levels (PPL) were regularly measured via venous blood draw, and subjective drug intensity (SDI) was assessed using a 0–10 scale.

#### Functional MRI acquisition

Participants were scanned on one of two Siemens 3T MAGNETOM Prisma scanners (n=15 on Scanner A; n=13 on Scanner B), with all scans for each participant acquired on the same scanner. Scanner A used a 64-channel head coil (TR=2000 ms, TE=30 ms, flip angle=90°, 64×64 matrix, 3.6×3.6 mm in-plane resolution, 32 slices, 3.0 mm thickness, 10 minutes, 300 volumes). Scanner B used a 32-channel head coil (TR=800 ms, TE=37 ms, flip angle=52°, 104×104 matrix, 2×2 mm in-plane resolution, 72 slices, 2.0 mm thickness, multiband factor=8, 10 minutes, 750 or 375 volumes).

#### Functional MRI Pre-processing

BOLD data were pre-processed using SPM12 and denoised in CONN. The pipeline included slice-timing correction (Scanner A only), realignment, unwarping, co-registration, segmentation, normalisation to MNI152 space, and smoothing (6 mm FWHM). Denoising included linear detrending, aCompCor regression (first five white matter and CSF components with derivatives), motion parameter regression (six parameters with derivatives), volume censoring (global signal z>4 or motion>2 mm), and bandpass filtering (0.008–0.09 Hz). Scanner B timeseries were downsampled to match Scanner A’s 2-second TR. For 25 participants fMRI data of suitable quality were available at the appropriate timepoints.

#### pcASL MRI acquisition

Scanner A used a 2D EPI gradient spin-echo sequence (TR=4000 ms, TE=12 ms, flip angle=90°, 64×64 matrix, 3×3 mm in-plane resolution, 20 slices, 5.0 mm thickness, label duration=1508 ms, single post-label delay=1500 ms, 40 control/label pairs, 5.33 minutes). Scanner B used a 3D turbo gradient spin-echo sequence (TR=4100 ms, TE=3.78 ms, flip angle=120°, 96×96 matrix, 2.5×2.5 mm in-plane resolution, 40 slices, 3.0 mm thickness, label duration=1508 ms, five post-label delays (500, 1000, 1500, 2000, and 2500 ms), 12 control/label pairs, 7.11 minutes). M0 calibration scans were acquired on both scanners.

#### pcASL MRI preprocessing

pcASL data were pre-processed and quantified using BASIL in FSL (https://fsl.fmrib.ox.ac.uk/fsl/docs/physiological/basil.html). The pipeline included realignment, distortion correction, label/control subtraction, and calibration to absolute CBF values (ml/100g/min) using M0 scans and single-compartment kinetic modeling. Images were normalized to MNI152 space and smoothed with an 8 mm FWHM kernel. Suitable quality pcASL data were available at the required time points for 25 participants.

### fMRI correlation metrics

Scripts for transforming C-PAC outputs to the appropriate input format and for performing each fMRI analysis are publicly available (https://github.com/Pneumaethylamine/dOccLS).

#### Atlas and region selection

Unless otherwise stated, all measures were calculated using a combined atlas of the Schaefer 200-region parcellation and the 32-region S2 Tian subcortical atlas (72). For network connectivity, graph theory, and information-entropy analyses, we extracted time series data using the Schaefer 200-parcel atlas, and correlations were estimated using Nilearn in C-PAC (73). Each Schaefer 200-region region was assigned to one of seventeen networks as described in the Yeo-17 network parcellation, which was used for estimating network-connectivity scores (74). These networks include: Control (A, B, C), Default Mode (A, B, C), Dorsal Attention (A, B), Limbic (A, B), Salience/Ventral Attention (A, B), Somatomotor (A, B), Temporal Parietal, Visual Central, and Visual Peripheral. Normalised Spatial Complexity and Sample Entropy used Yeo-17 Networks as seeds.

#### Global correlation (GCOR)

Pearson’s correlation between the time series for each pair of regions was calculated to yield a whole-brain connectivity matrix. The regional GCOR score was calculated as the row-mean, excluding the diagonal.

#### Atlas-based network connectivity

Network connectivity was quantified for both between-network and within-network connections. Let C(rₐ, rᵦ) denote the functional connectivity score between regions rₐ and rᵦ. The connectivity Aᵢ□ between networks Nᵢ and N⍰ was computed as: Aᵢ□ = median(C(rₐ, rᵦ)) for all rₐ ∈ Nᵢ and rᵦ ∈ N⍰ when rₐ ≠ rᵦ.

### fMRI information entropy metrics

Information-entropy metrics were calculated using the in-house developed Copenhagen Brain Entropy (CopBET) toolbox, available on GitHub https://github.com/anders-s-olsen/CopBET. For more detailed methods, please see (71). Each applied metric is summarised briefly below. The measures Geodesic distribution entropy, Dynamic Conditional Correlation and Normalised Global Spatial Complexity apply a Shannon entropy quantification 𝐻(𝑋) = − ∑𝑥∈𝑋 𝑝(𝑥) ⋅ 𝑙𝑜𝑔2𝑝(𝑥) where H(X) refers to the Shannon entropy of probability mass function X containing bins (x) with height (p).

#### Geodesic distribution entropy

Geodesic distribution entropy is a measure of the entropy of static connectivity given by the matrix of Pearson correlation coefficients, R, calculated as described above (75). Degree refers to the number of non-zero elements in any given row of a thresholded matrix. ROI-specific degrees are computed based on R, with 𝑁=200, using absolute correlation values. The goal of the thresholding for this analysis is to reach a pre-specified mean degree across rows. To achieve this, a threshold below which all absolute values are set to 0 is gradually increased until the mean number of non-zero elements is at the desired level. Here, we applied a scan-specific threshold that produced a mean degree range of 20-40. We report in the main text only the results from a mean degree of 40, as this aligns with a threshold applied in the previous literature. This means that each scan may have a different absolute threshold value, but nearly identical mean degree. The matrix was then binarised, setting all non-zero elements to 1. The “shortest path length” was then computed as the fewest edges one must traverse to go from one node to another using the MATLAB function “distances”. The Shannon entropy of the distribution of path lengths from each node to all other nodes was then calculated.

#### Dynamic conditional correlation entropy

Windowless framewise correlation coefficients were calculated for all edges using the Dynamic Conditional Correlation (DCC) toolbox (76,77). Subsequently, the probability distribution over each ROI-to-ROI DCC time series was established, and the Shannon entropy was calculated. Each ROI was assigned to one of the Yeo-17 networks. Each ROI-to-ROI pair was then assigned to its respective network-to-network association (e.g., motor-motor, default mode-motor, etc.); the mean entropy of each network-to-network association was calculated.

#### Normalised spatial complexity (NSC)

NSC entropy was calculated using principal component analysis (PCA) of the voxelwise fMRI time series across the whole cortex or each Yeo-17 Network separately (78). This yields a set of eigenvalues. Let λ₁, λ₂, …, λ□ be the eigenvalues obtained from the PCA, where m is the total number of eigenvalues. These eigenvalues are first normalised by dividing each by the sum of all eigenvalues: 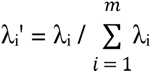, where λᵢ’ is the normalised eigenvalue and the sum is taken over all i from 1 to m. The NSC entropy is then computed as: 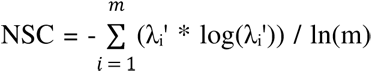, where the sum is taken over all i from 1 to m. The division by ln(m) serves to normalise the entropy value, ensuring it falls between 0 and 1. This calculation is performed both on the whole-brain level and for each region of interest defined by the atlas, resulting in a global NSC entropy value and a set of regional NSC entropy values for each fMRI session.

#### Multi-scale sample entropy

Sample entropy is defined as the negative logarithm of the conditional probability that if two vectors with length m (set to 2) are dissimilar below a threshold distance r (set as 0.3), then vector pairs with length m+1 will also have distance below the threshold. Sample entropy for scales 1-5 was evaluated for each network, meaning that each time-series was split into non-overlapping windows of length (scale) s volumes and the means of each window were concatenated to form a condensed time-series upon which sample entropy was calculated (79). Sample entropy was calculated for each voxel in the Yeo-17 networks and then averaged across voxels to return a score for each scale for each network. Scale 1 (i.e., no downsampling) is reported in the main text; all scales are reported in the Supplementary Table S5.

#### Lempel-Ziv complexity

The BOLD time series for each ROI were first Hilbert-transformed. The amplitude of the Hilbert series was then binarised around the mean amplitude for that region, i.e., assigned as 1 if greater than the mean and 0 if less. These binarised time series were combined into a T×N matrix, where T is the number of time points and N = 232 is the number of regions. This matrix was collapsed into a single vector to compute the Lempel-Ziv complexity over time (LZct, LZ78 algorithm) wherein regional time series were concatenated. LZct represents a calculation of the temporal entropy of each ROI (80,81).

### fMRI graph theory metrics

Graph theory metrics used functions from the Brain Connectivity Toolbox (2019 March version) (82).

#### Modularity

Modularity was calculated by applying the community_louvain function from the Brain Connectivity Toolbox (BCT) (82) with γ = 1 and asymmetric treatment of negative weights to the weighted, undirected Pearson’s correlation matrix for each scan individually. This was repeated 100 times, and the maximum value was saved as the absolute modularity. Subsequently, 100 null graphs were created using the null_model_und_sign function in BCT; for each null graph, the same procedure for calculating modularity was applied, i.e., taking the maximum of 100 repetitions of applying the community_louvain function. The observed modularity was normalised with respect to the mean modularity of these null models, i.e., normalised_modularity = absolute_modularity / null_modularity.

#### Small-worldness

A positive-only matrix was used by removing all negative correlations because small-worldness stems from calculating path-length, which is ill-described for graphs with negative correlations. Further, we did not consider it sensible to take the absolute values, as negative correlations represent distinct neural phenomena from positive correlations, and there is no consensus strategy for how to account for these in the estimation of small-worldness (83).

First, the mean clustering coefficient (C) across all regions was calculated using the BCT function clustering_coef_wu. The average path length (L) was calculated using the BCT function distance_wei_floyd(‘log’) as the mean of the non-infinite, non-zero path lengths. We generated 100 null graphs using the BCT function null_model_und_sign on the unthresholded matrix, which was then thresholded to include only positive edges. C and L were calculated for each null graph, and observed small-worldness was normalised as SM_norm_ = (C / mean(C_null_)) / (L / mean(L_null_)), where C_null_ and L_null_ are C and L, respectively, calculated on null graphs.

### Multimodal analysis

#### Registration and regional alignment

To enable cross-modal comparisons between CBF and BOLD fMRI (GCOR), we quantified both modalities within the same set of brain regions defined by the AAL3 atlas (68). CBF data were already processed within the AAL regions. To ensure spatial correspondence, individual voxel-wise GCOR maps were transformed into AAL3 space, and regional mean values were extracted from the same atlas-defined regions. For PET data, we used previously computed neocortical 5-HT2AR occupancy estimates for each participant.

#### Cross-modality correlation analysis

We used Spearman’s rank correlation (ρ) to assess the relation between neocortical 5-HT2AR occupancy and changes in regional CBF, between occupancy and changes in regional GCOR, and between changes in CBF and GCOR. Changes for CBF and GCOR scores were calculated as LSD minus baseline. Correlations were run across participants for each region to identify spatial variability in how receptor occupancy relates to functional brain changes.

### Statistics

Given the densely sampled but limited cohort (n = 7) and heterogeneous dosing regimens, we prioritised effect size analyses to characterize LSD-induced hemodynamic and functional connectivity alterations. Effect sizes provide meaningful information about the magnitude and practical meaningfulness of the observed changes, particularly valuable in studies with smaller samples (84). While we initially explored linear mixed-effect models to account for repeated measures, these often failed to converge or produced singular fits. Therefore, to ensure consistent modeling across measures, we averaged across densely sampled baseline and LSD runs to calculate participant- and session-specific maps. We computed Cohen’s dz values as the mean difference between conditions (LSD - baseline) divided by the standard deviation of those differences. 95% confidence intervals on the effect size are calculated as dz ± 1.96 * (sqrt((1/n) + (dz^2 / (2*n))) where n = 7. For CBF regional effects, we supplemented the effect size analyses with a step-down permutation procedure that leveraged all possible permutations of our dataset to control for multiple comparisons (see Supplementary Figure S7 and Supplementary Table S2 for details).

For the relation between average subjective drug intensity and occupancy, a sigmoid function was fit where the parameters a, b, and c were estimated using nonlinear least squares regression on the model data. The sigmoid model was implemented in R (v4.3.1) using the following code:

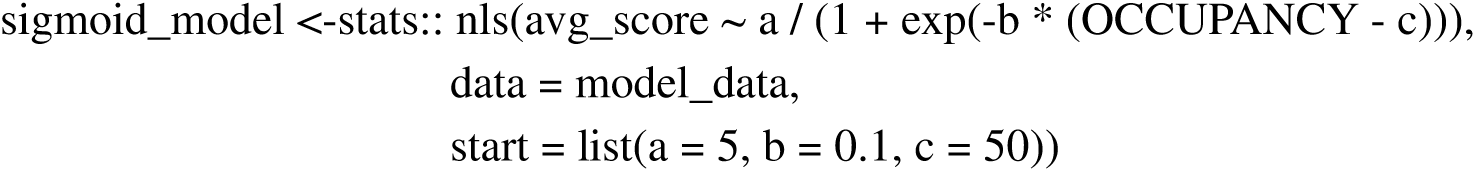

#### Plasma LSD quantification

Plasma samples (100 µL) were spiked with 20 µL internal standard (LSD-d3) and protein precipitated with acetonitrile (700 µL) using a fully automated robotic system. The supernatant (700 µL) was acidified with 50 µL 10% formic acid in acetonitrile, evaporated, and reconstituted in 100 µL 12.5% methanol:12.5% acetonitrile in 0.05% formic acid.

LSD was quantified by ultra-high-performance liquid chromatography–tandem mass spectrometry (UHPLC–MS/MS) using a Kinetex Biphenyl column (2.6 µm, 2.1 x 50 mm), and gradient elution of A: 1 mM ammonium formate in 0.1% formic acid and B: 1:1 methanol:acetonitrile with 0.1% formic acid. The column temperature was held constant at 50°C, the flow rate was 0.50 mL/min, and the total run time was 3.9 min. The injection volume was 2 µL. The quantification was performed using multiple reaction monitoring (MRM) in positive ionization mode. The two MRM transitions used for LSD were m/z 324 -> 223 and 324 -> 207. The MRM transition used for the internal standard LSD-d3 was m/z 327 -> 226.

Five calibrators in plasma were freshly prepared for each series, and two quality control (QC) samples in plasma were included in each series in duplicates. The measurement range was 0.0001-0.01 mg/kg (0.28 - 28.5 nM) using a linear calibration model and a weighting factor of 1/x. The method showed acceptable accuracy and precision for the QCs according to international guidelines.

Concentrations were quantified in mcg of LSD per kg blood, which were converted to nanomolar values using an estimated plasma density of 1.02 g/ml and the molecular weight of LSD of 323.43 g/mol.

### *CYP2D6* genotyping

*CYP2D6* genotyping was performed according to established methods at the Danish Epilepsy Centre Laboratory Unit using validated real-time PCR assays (For detailed methodology, see Jensen et al.) (85).

## Code availability

The code used for analyses are available on Github https://github.com/Pneumaethylamine/dOccLS.

## Data availability

Data from this study are available through the Center for Integrated Molecular Brain Imaging (Cimbi) Database and Biobank at Rigshospitalet, Copenhagen (86). Researchers can request access to the dataset by completing a standardised application form available at the Cimbi website (https://nru.dk/index.php/research-menu/research-groups/115-the-cimbi-database-and-biobank) (86)

## Author contributions

D.E.W.M. and K.L. contributed equally to this work as co-first authors. D.E.W.M. designed the study, developed protocols, collected data, performed data analysis, created visualizations, and wrote the manuscript. K.L. collected data, performed data analysis, created visualizations, and wrote the manuscript. A.J., K.H.R.J., and C.H.N. provided medical supervision throughout study implementation. F.H. provided supervision and assisted with regulatory submissions. N.F., V.N., E.S., P.S.A., and A.S. recruited and screened participants and assisted with data collection. M.G. and P.P.R. provided psychological support during the psychedelic sessions. P.S.J. managed the database and data organization. V.S. performed radiochemistry for the PET tracers. S.S.J. and M.K.K.N. conducted quantification of plasma drug levels. T.L.A. supervised PET and MRI scanning procedures. D.S.S. provided psychology supervision and guidance. C.S. supervised PET data analysis. P.M.F. and G.M.K. contributed equally to this work as co-senior authors. They designed the study, supervised the project implementation, provided resources, and contributed to manuscript writing and revision. All authors reviewed and approved the final version of the manuscript.

## Acknowledgements

We would like to thank Dr. Cedric Hysek and Dr. Matthias Liechti for supplying LSD for the study, Lone Freyr and Emilie Henriksen for facilitating and planning all of the scans. Dorthe Givard for her endlessly patient administrative support. Prof. Brice Ozenne for patient statistical advice. Dr. Maja Marstrand-Jørgensen for helping with setting up the scan parameters and REDcap databases, and for looking after our baby daughter during the long scan days. The John and Birthe Meyer Foundation is gratefully acknowledged for the donation of the Cyclotron and PET-MRI scanner.

## Supplementary materials

### Supplementary Results

- Adverse effects
- LSD effects on emotional response to music

### Supplementary Figures

- S1. Relation between CYP2D6 metabolizer status and LSD plasma levels
- S2. Relation between weight-adjusted LSD dose and peak plasma concentrations
- S3. In-scanner comfort ratings during scans
- S4. Dose-occupancy relation for LSD
- S5. LSD-induced changes in cerebral blood flow and internal carotid artery flow
- S6. Regional cerebral blood flow measurements at baseline and after LSD administration
- S7. Distribution of t-statistics for LSD effects on cerebral blood flow across brain regions
- S8. Individual changes in cerebral blood flow following LSD administration.
- S9. Individual changes in global brain connectivity following LSD administration
- S10. Study protocol overview and participant flow (CONSORT diagram)
- S11. Schematic illustration of the study design and participant progression through different scanning phases.
- S12. Quality control pipeline across imaging modalities
- S13. Music-evoked emotional responses measured by the Geneva Emotional Music Scale (GEMS)
- S14. Hysteresis curves showing the relation between plasma psilocybin concentration and subjective drug intensity
- S15. Regional correlations between cerebral blood flow, global correlation, and receptor occupancy

### Supplementary Tables

- S1. Participant characteristics and PET scan parameters across study phases
- S2. Regional cerebral blood flow, internal carotid artery diameter, and flow measures
- S3. Global correlation (GCOR) regional effects with and without GSR
- S4. Other functional MRI results with and without GSR
- S5. AAL3 regions combined to create regions of interest

## Adverse effects

Three participants experienced transient adverse events. One participant reported mild nausea, dizziness, and moderate anxiety with ego dissolution, which resolved concurrently with the acute drug effects. Another participant experienced severe emesis and nausea that resolved with drug clearance, followed by temporary difficulty with distance visual accommodation that spontaneously resolved within 72 hours. A third participant reported acute anxiety associated with the scanner environment that dissipated upon exiting the scanner, and mild sleep disturbances lasting approximately one week post-administration, with spontaneous resolution. No serious adverse events occurred, and no medical intervention was required for any participant.

## LSD effects on emotional response to music

LSD enhanced emotional responses to music during the scan compared to the baseline scan (Supplementary Figure S14). The most pronounced effect was observed in the item ‘Transcendence’, showing a large increase (dz = 1.63) under LSD compared to baseline. Moderate increases were also observed in Strength (dz = 0.89) and Joy (dz = 0.71). Smaller but notable effects were seen in Wonder (dz = 0.55), Tension (dz = 0.47), and Tenderness (dz = 0.46). Sadness showed a modest increase (dz = 0.40), whereas Nostalgia and Peacefulness demonstrated the smallest effects (both dz = 0.26).

---

**Supplementary Figure S1.**
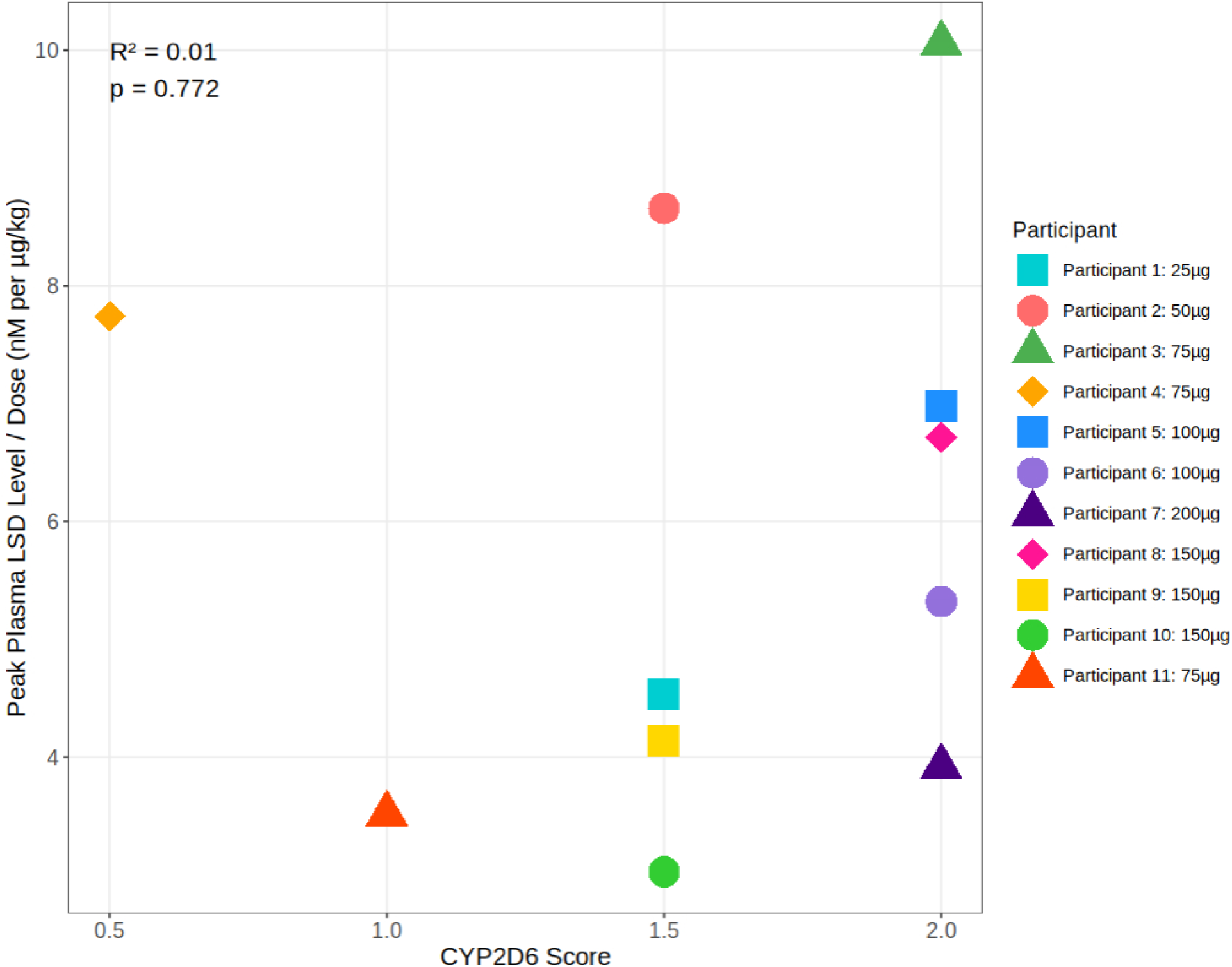
| Relation between CYP2D6 metabolizer status and LSD plasma levels. Peak plasma LSD concentrations normalised by body weight-adjusted dose (nM per µg/kg) plotted against CYP2D6 metabolizer scores for all participants (n=11). Each point represents an individual participant, with different shapes and colors identifying unique participants. *CYP2D6* scores range from 0.5 (poor metabolizer) to 2.0 (normal metabolizer).

**Supplementary Figure S2.**
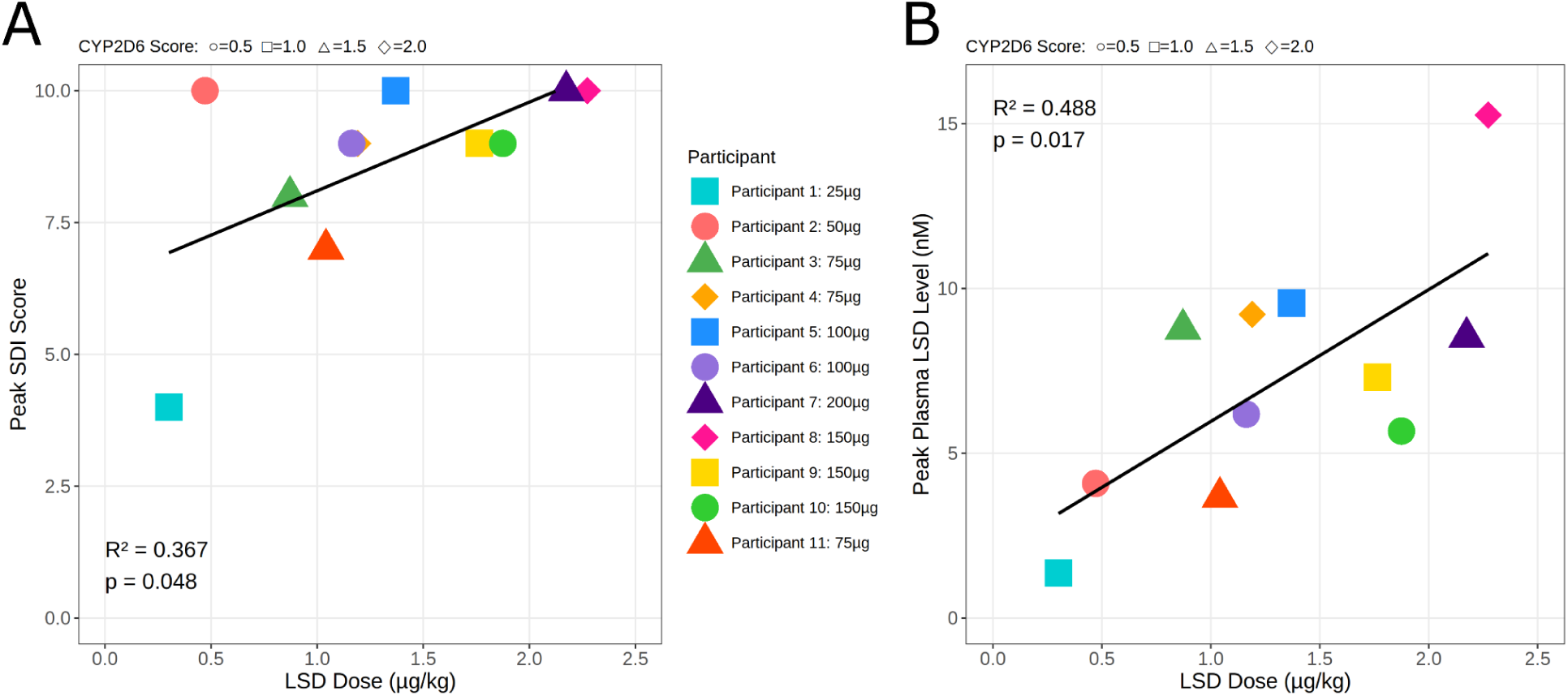
| Relation between weight-adjusted LSD dose and peak plasma concentrations. A, Peak subjective drug intensity plotted against body weight-adjusted LSD dose (μg/kg) across study participants (n = 11). The black line shows the linear regression (R²=0.37, p=0.048), demonstrating a significant positive correlation between administered dose and peak subjective drug effects. B, Peak plasma LSD concentrations plotted against body weight-adjusted dose (μg/kg) across study participants (n=11). Individual CYP2D6 metaboliser scores are indicated by marker shapes (circle=0.5, square=1.0, triangle=1.5, diamond=2.0), reflecting varying metabolic capacity. The black line shows the linear regression (R²=0.49, p=0.017), demonstrating a significant positive correlation between administered dose and peak plasma levels. Peak weight-adjusted plasma LSD levels showed little correlation with CYP2D6 metabolizer status (R² = 0.01, P =0.78).

**Supplementary Figure S3.**
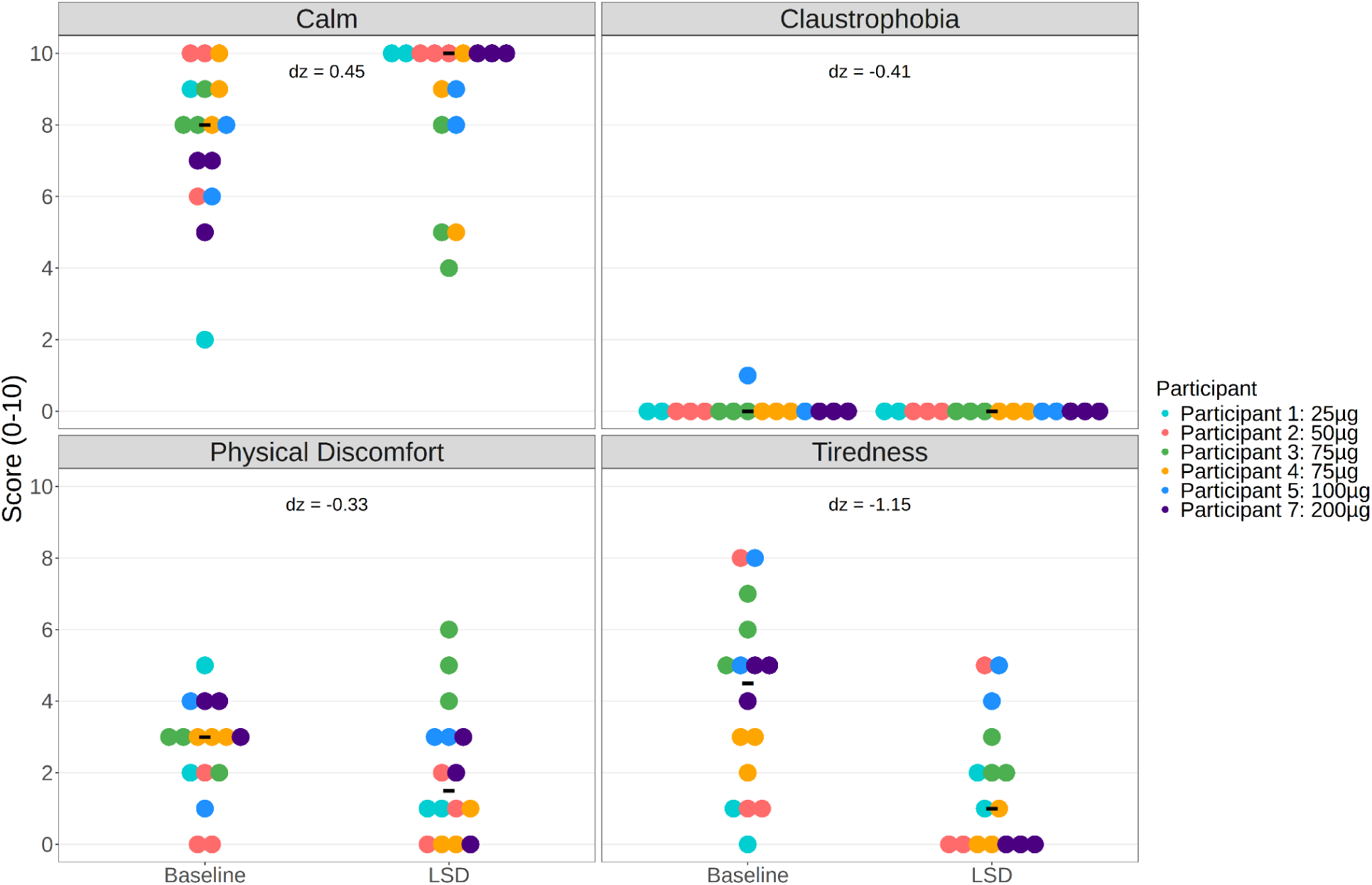
| In-scanner comfort ratings during scans. Self-reports during baseline and LSD conditions (n=6). Effect sizes (Cohen’s dz) show reduced Tiredness (dz = 1.15) with minimal changes in Calm, Claustrophobia, and Physical Discomfort ratings. Black lines represent the median value. Each color represents a unique participant receiving doses ranging from 25-200 µg of LSD. Ratings were collected immediately following each scanning sequence during peak drug effects, approximately 2-3 hours post-administration.

**Supplementary Figure S4.**
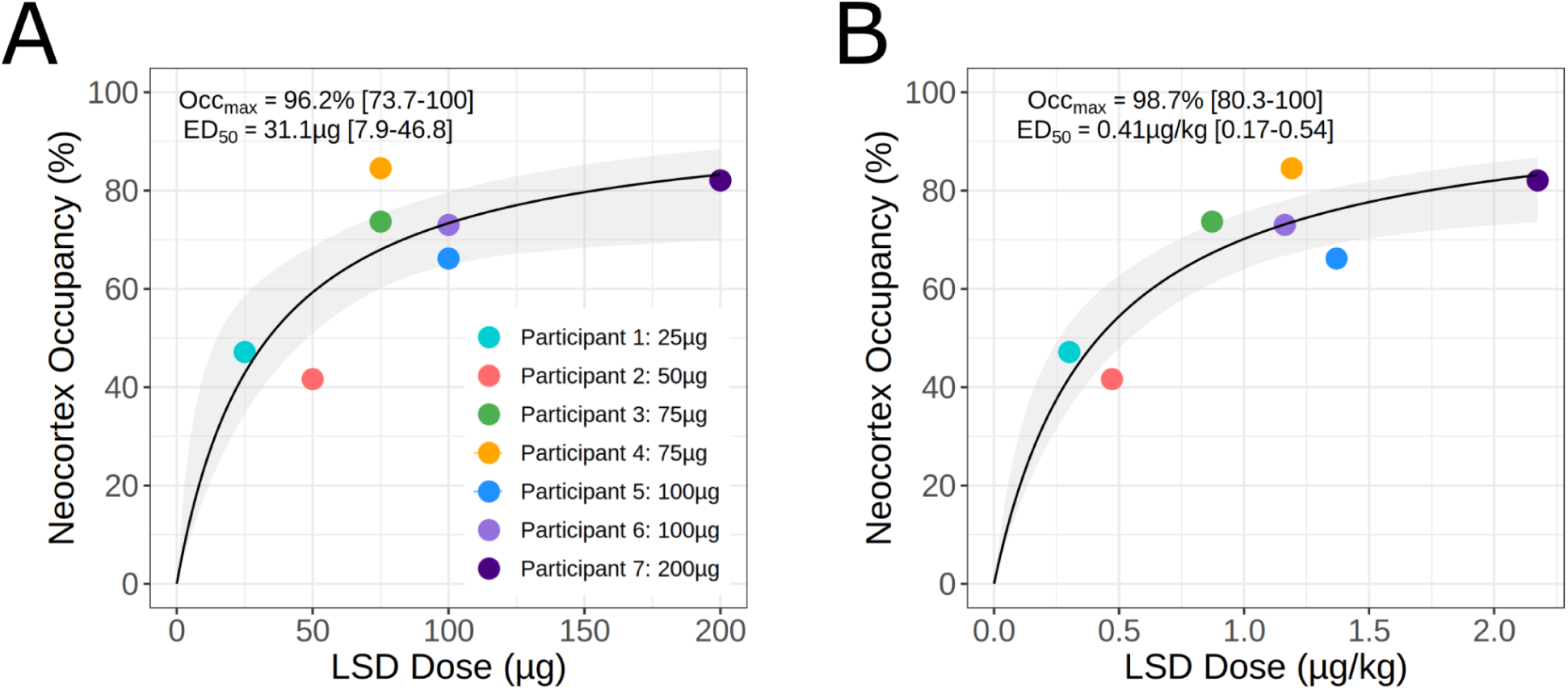
| Dose-occupancy relation for LSD. A: LSD dose (µg) versus receptor occupancy fitted with Hill-Langmuir model. The solid black line shows the model fit, and bootstrap-derived 95% confidence intervals are shown in grey shading. Model estimates from the original fit with bootstrap confidence intervals indicate maximum occupancy of 96.2% [73.5-100%] and ED_50_ of 31.1 µg [7.6-46.4 µg]. **B:** Body weight-adjusted LSD dose (µg/kg) versus receptor occupancy fitted with Hill-Langmuir model. The solid black line shows the model fit with bootstrap-derived 95% confidence intervals in grey shading. Model estimates indicate maximum occupancy of 98.7% [80.3-100%] and ED_50_ of 0.41 µg/kg [0.17-0.54 µg/kg]. The improved coefficient of determination (R² = 0.75 versus R² = 0.58 for unadjusted dose) suggests that body weight adjustment better predicts receptor occupancy.

**Supplementary Figure S5.**
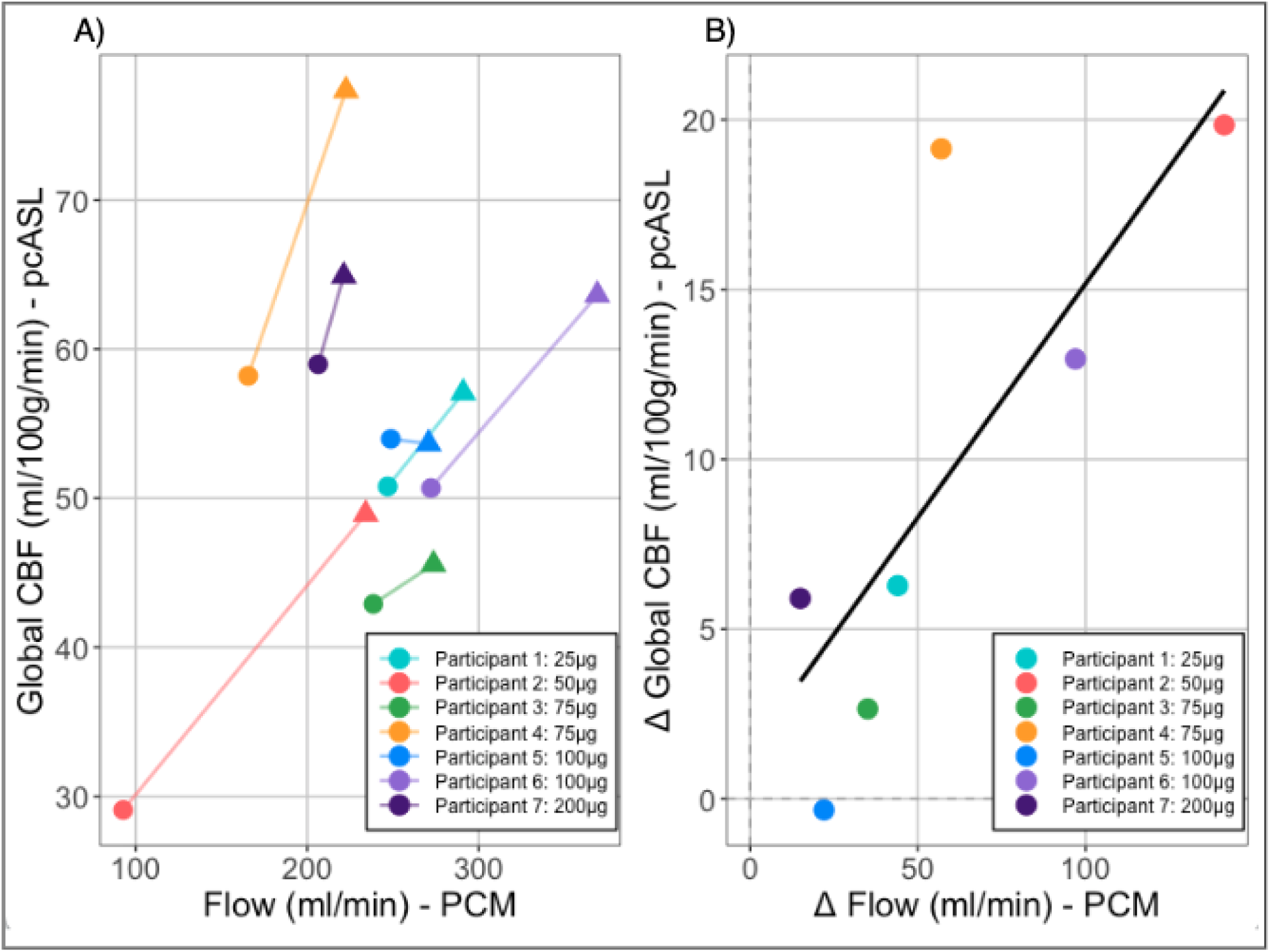
| LSD-induced change in cerebral blood flow and internal carotid artery flow. A) Global cerebral blood flow (CBF) measured using pseudo-continuous Arterial Spin Labeling (pcASL) plotted against internal carotid artery flow measured with Phase Contrast Mapping (PCM), with circles representing baseline measurements and triangles representing post-LSD administration for seven participants at varying doses (25-200μg). B) Relation between LSD-induced changes (Δ) in global CBF and changes in internal carotid artery flow, showing a significant positive correlation (r = 0.786, p = 0.036).

**Supplementary Figure S6.**
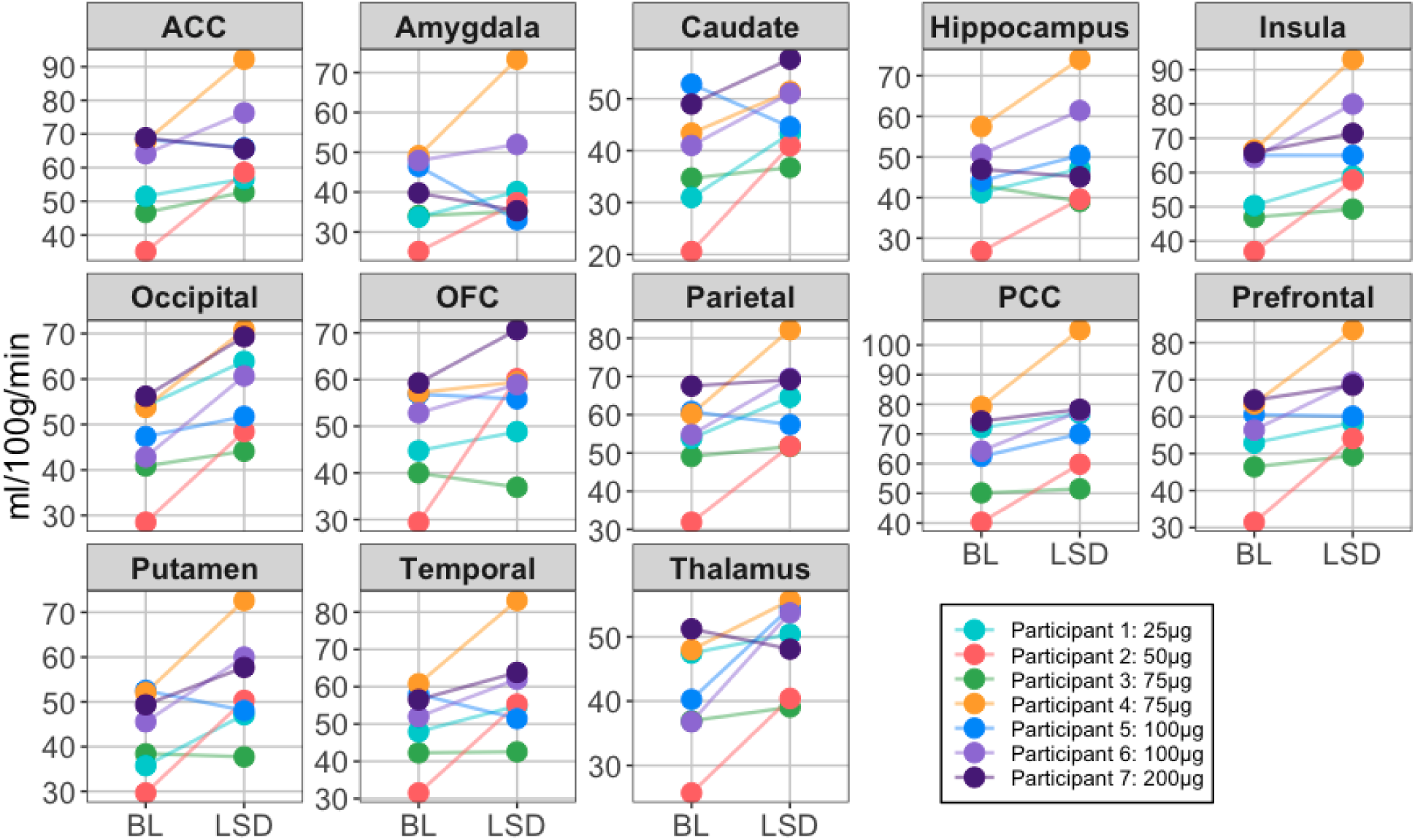
| Regional cerebral blood flow (CBF) measurements at baseline (BL) and after LSD administration across 13 brain regions. Each colored line represents a different participant who received varying doses of LSD (25-200µg), with measurements shown in ml/100g/min. Brain regions analysed include the anterior cingulate cortex (ACC), amygdala, caudate, hippocampus, insula, occipital cortex, orbitofrontal cortex (OFC), parietal cortex, posterior cingulate cortex (PCC), prefrontal cortex, putamen, temporal cortex, and thalamus.

**Supplementary Figure S7.**
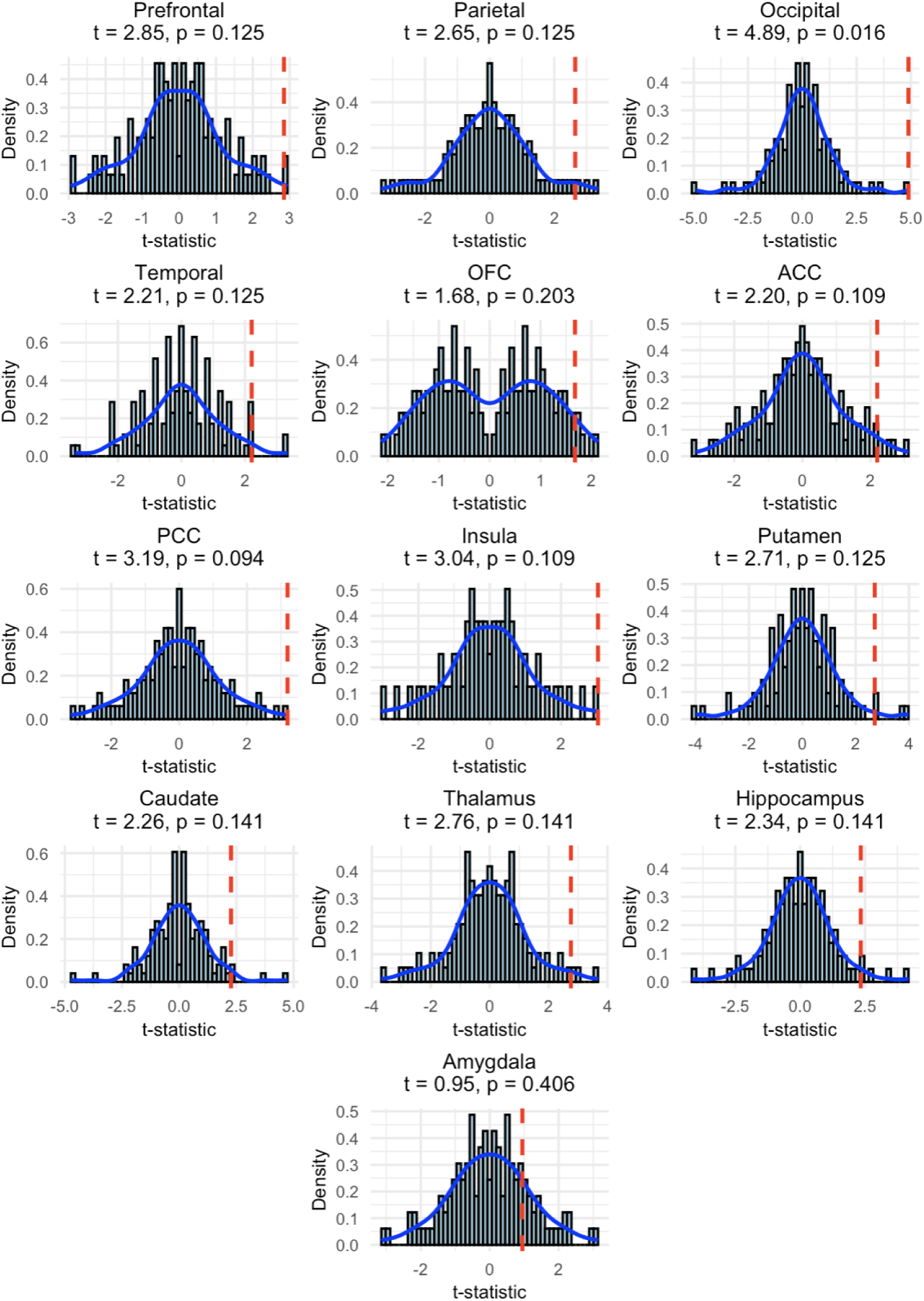
| Distribution of t-statistics for the LSD effect on cerebral blood flow across 13 brain regions. Each subplot shows the null distribution of t-statistics generated from permutation testing, represented by histograms (light blue bars) with overlaid density curves (blue lines). The observed t-statistic for each region is indicated by a vertical red dashed line. The test statistics (t-value) and stepdown-adjusted p-values are shown above each plot. The Occipital region shows the strongest effect (t = 4.89, p = 0.016), whereas the Amygdala shows the weakest effect (t = 0.95, p = 0.406).

**Supplementary Figure S8.**
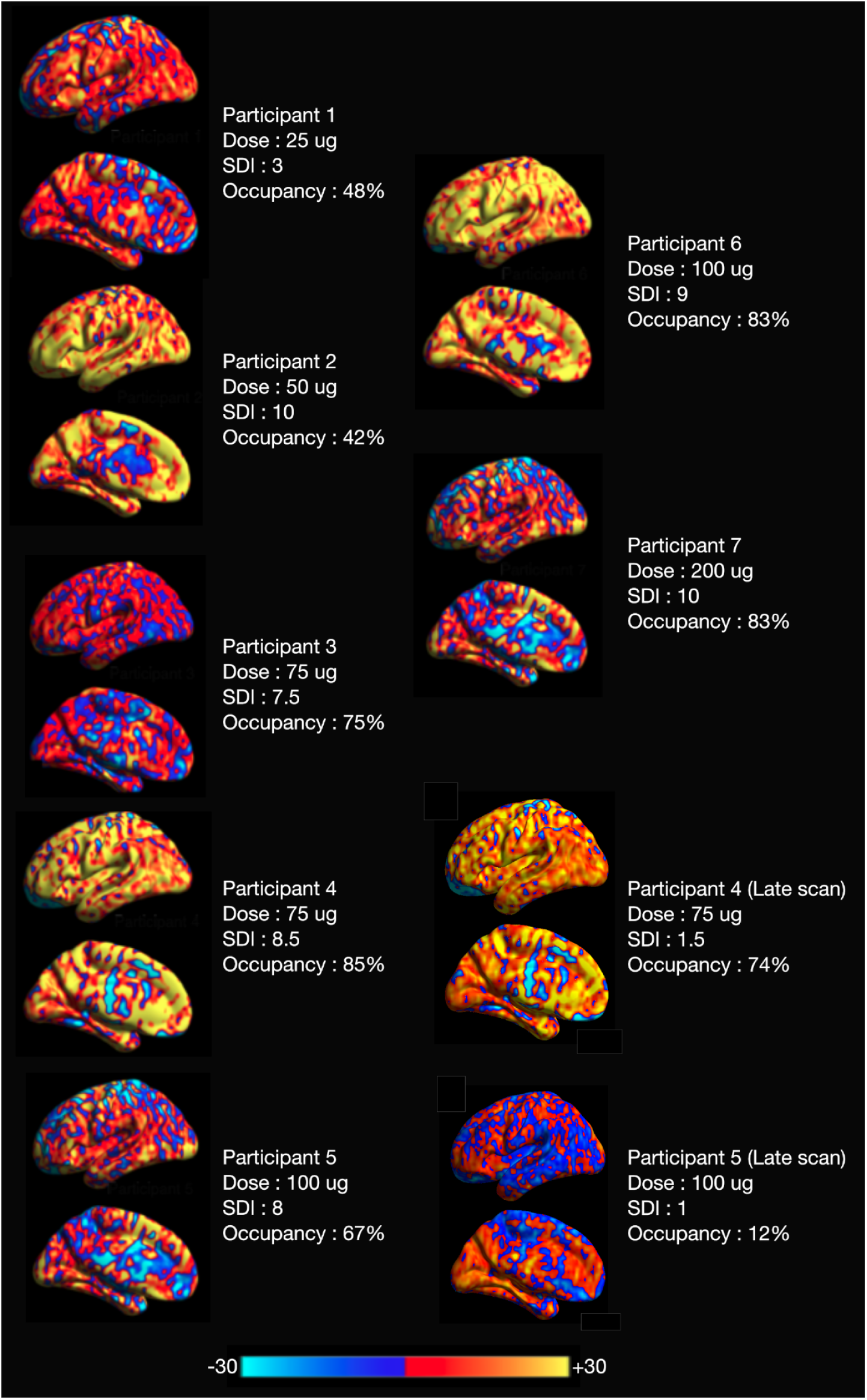
| Individual changes in cerebral blood flow following LSD administration. Changes in cerebral blood flow (CBF) for each participant following LSD administration compared to baseline, shown on inflated cortical surfaces. The color scale indicates CBF changes from -30 to +30 ml/100g/min, with red-yellow showing increases and blue showing decreases. Made using BrainNetViewer.

**Supplementary Figure S9.**
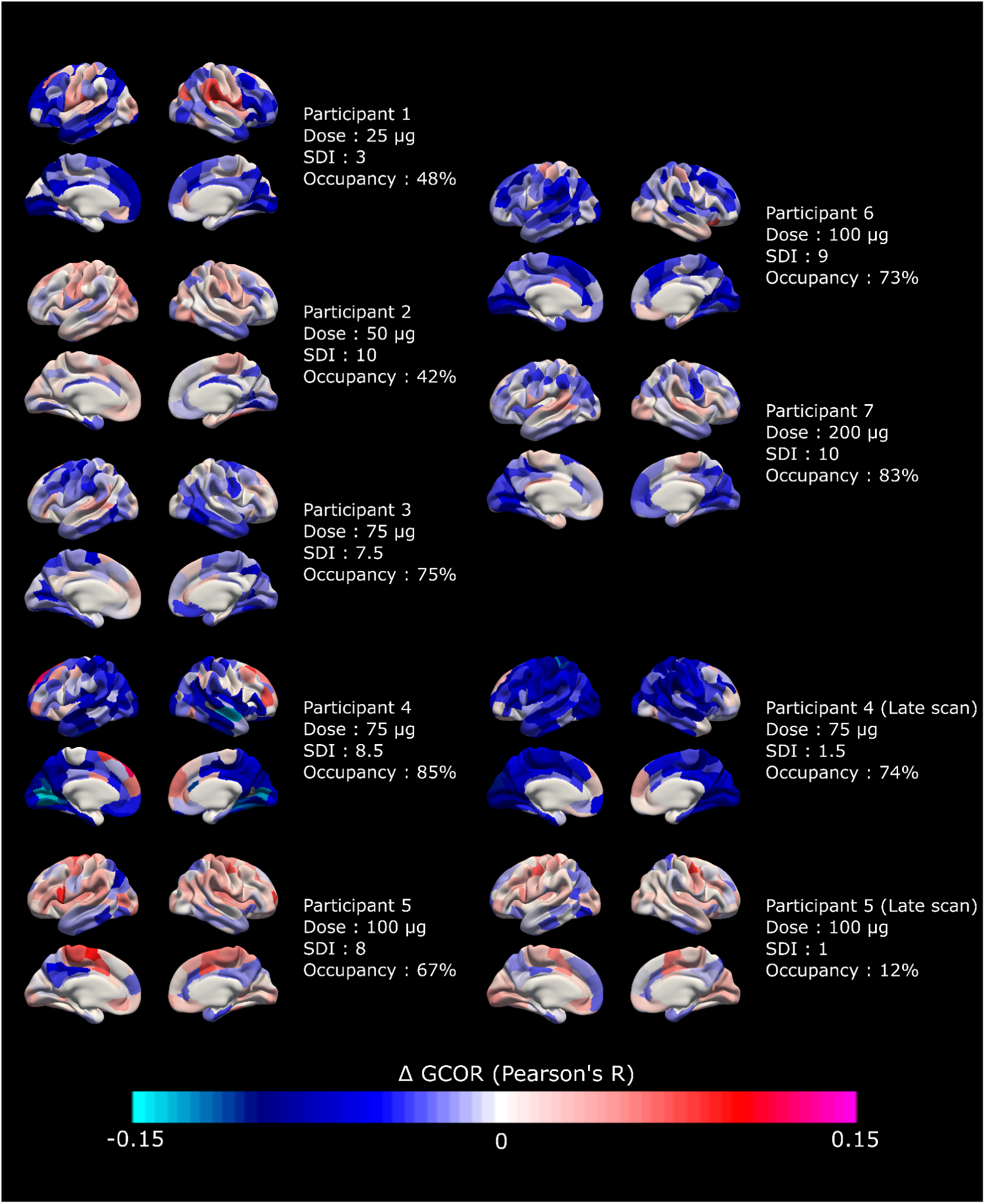
| Individual changes in global brain connectivity following LSD administration. Brain surface renderings showing changes in global functional connectivity for each of the seven participants included in the final analysis. Each panel displays lateral and medial views of both hemispheres with changes in connectivity represented on inflated cortical surfaces. Red colors indicate increases in global connectivity, whereas blue colors indicate decreases relative to baseline. Color intensity corresponds to the magnitude of connectivity change. All maps were generated using Freeview. Consistent patterns of decreased connectivity (blue) can be observed across visual and attention networks in most subjects, aligning with the group-level findings reported in the main text. SDI; Subjective Drug Intensity.

**Supplementary Figure S10.**
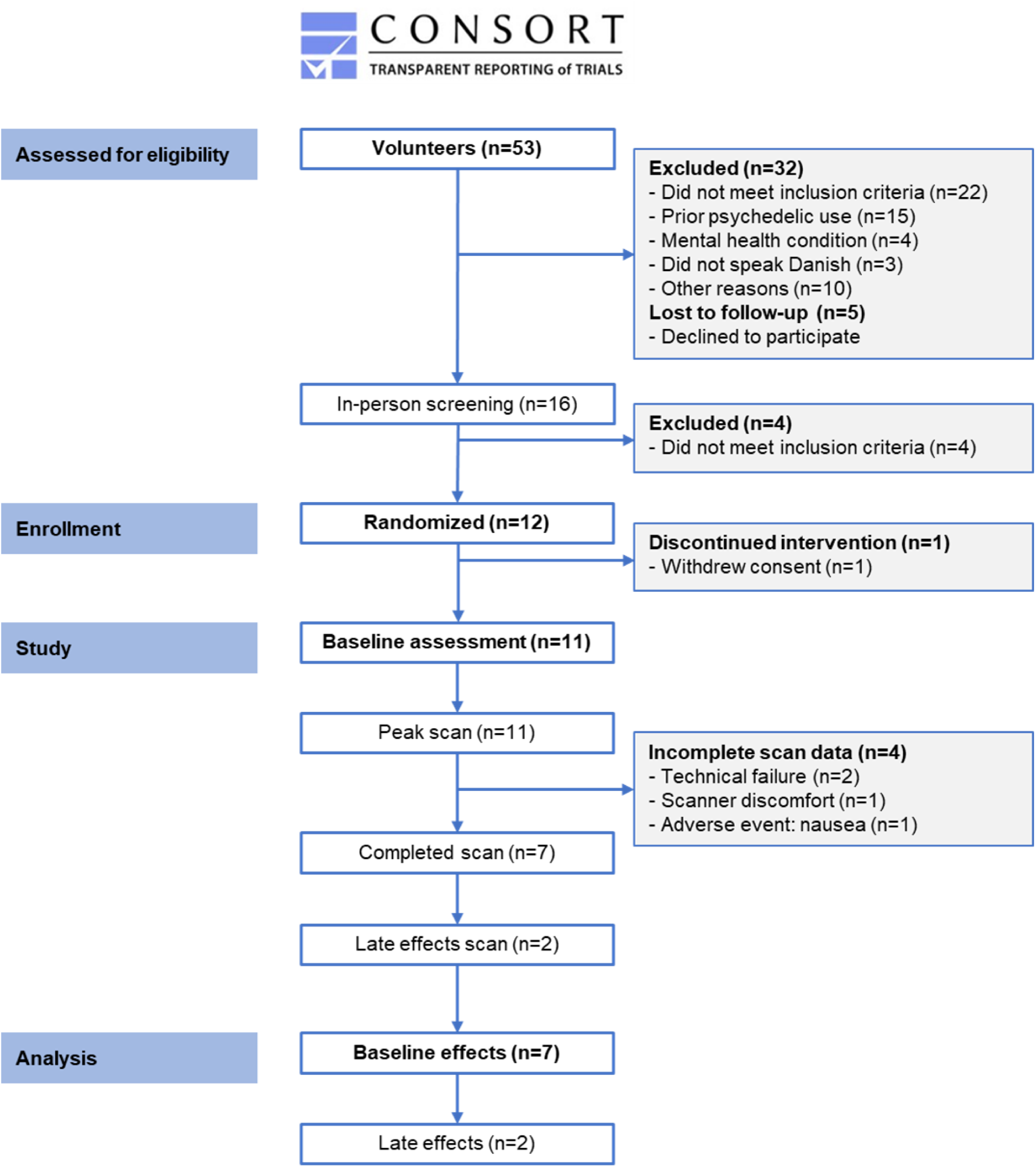
| CONSORT flow diagram showing participant recruitment and retention through the study.

**Supplementary Figure S11.**
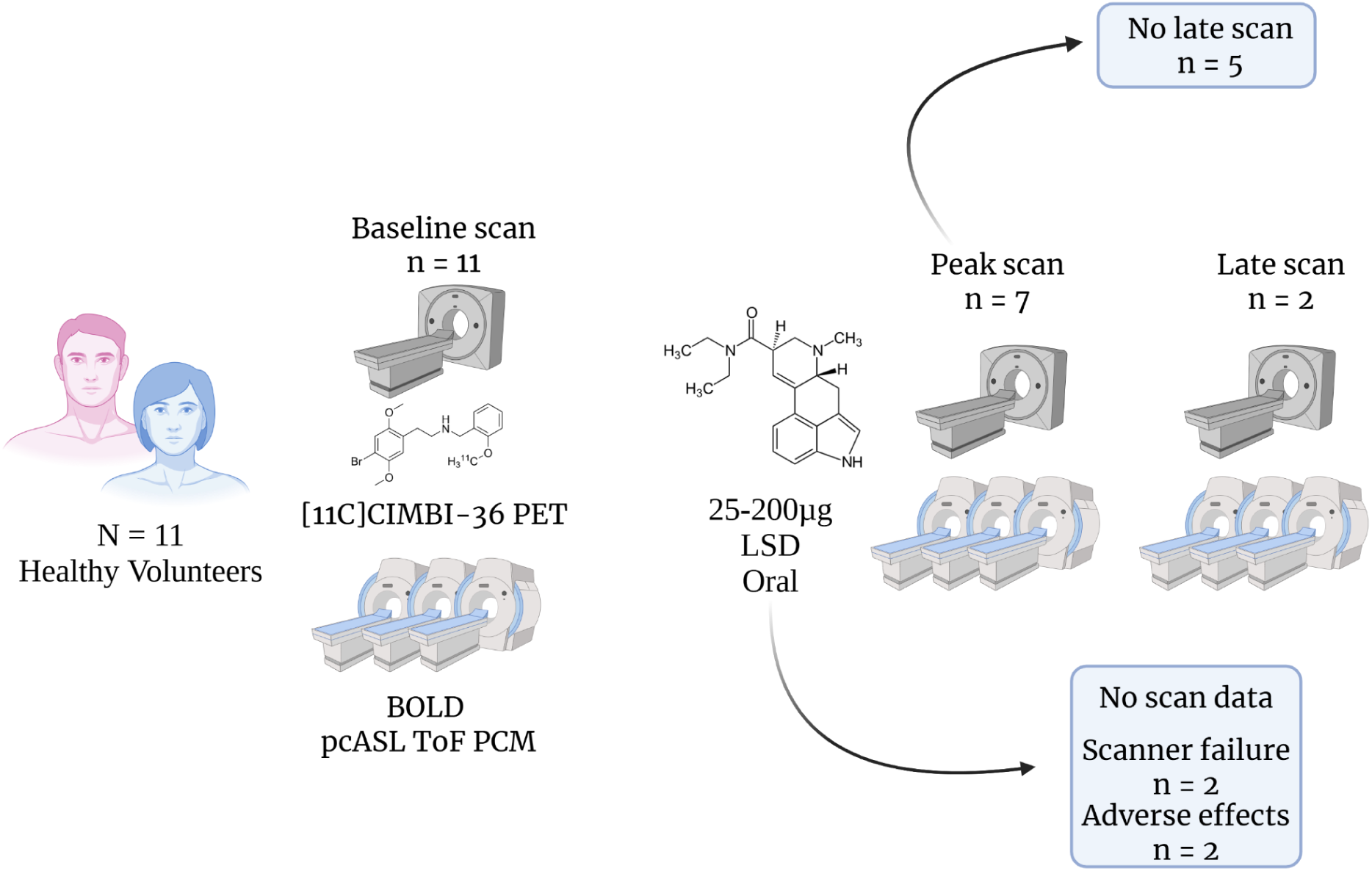
| Schematic illustration of the study design and participant progression through different scanning phases. All participants (n=11) underwent a baseline scan including [^11^C]CIMBI-36 PET imaging and multimodal MRI (multi-band multi-echo BOLD, pseudo-continuous arterial spin labeling, time-of-flight angiography, and phase contrast mapping). Following LSD administration (25-200μg oral), participants completed peak scans (n=7), and two of them also completed late scans. Four participants did not complete LSD scans due to scanner failure (n=2) or adverse effects (n=2). Of the seven completing peak scans, two completed late scans.

**Supplementary Figure S12.**
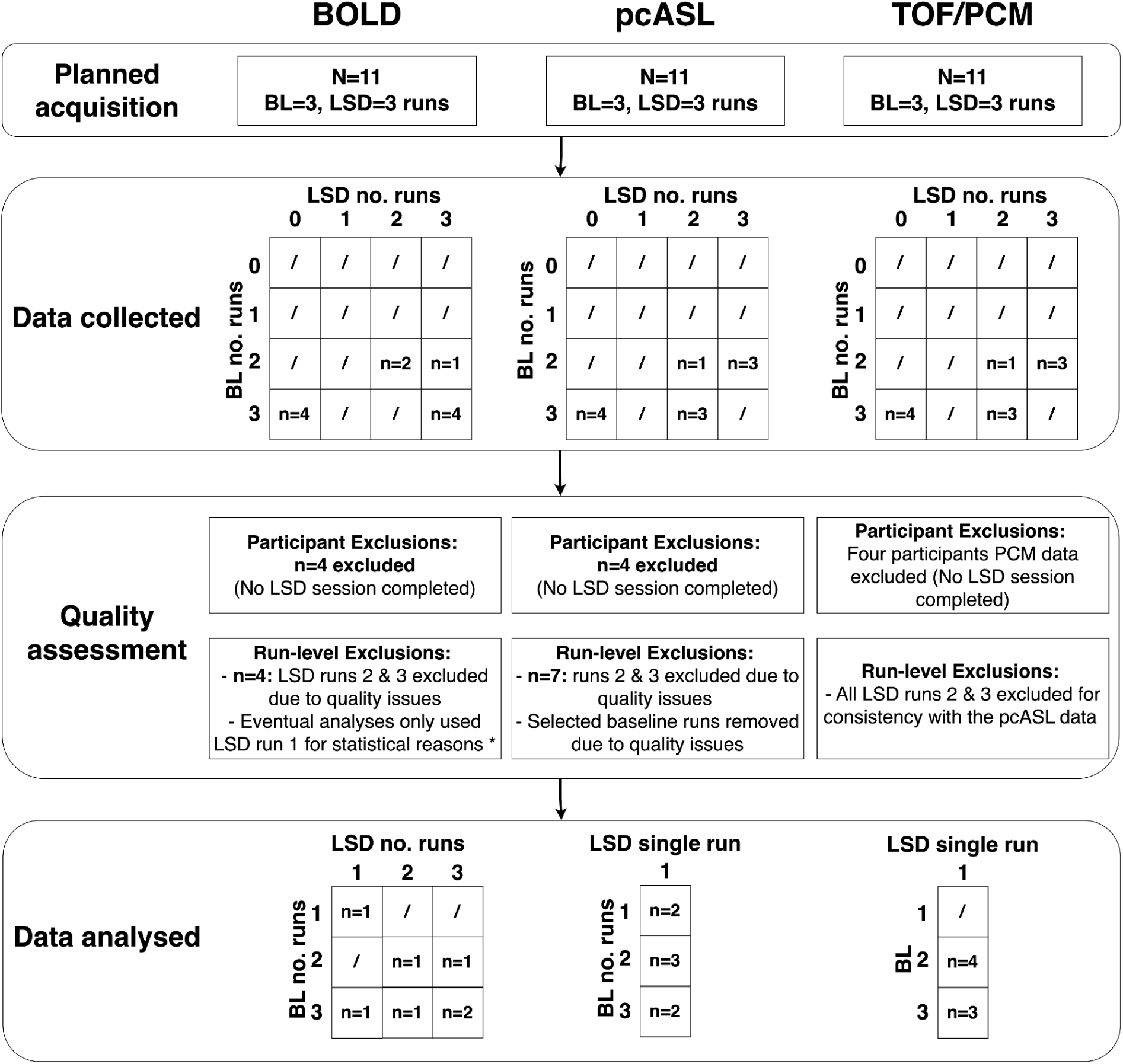
| Quality control pipeline across imaging modalities. The figure illustrates the quality control and data processing pipeline across the MRI modalities: BOLD (blood oxygen level dependent), pcASL (pseudo-continuous arterial spin labelling), and TOF/PCM (Time-of-flight, Phase contrast mapping). The planned acquisition for all modalities included BL=3 (Baseline) and LSD=3 runs. The “Data collected” matrices show the distribution of participants according to the number of baseline runs (y-axis, 0-3) and LSD runs (x-axis, 0-3) that were successfully collected. Forward slashes (/) indicate that no participants had that particular combination of runs. The “Data analysed” panel shows the final distribution of included runs for each modality.

**Supplementary Figure S13.**
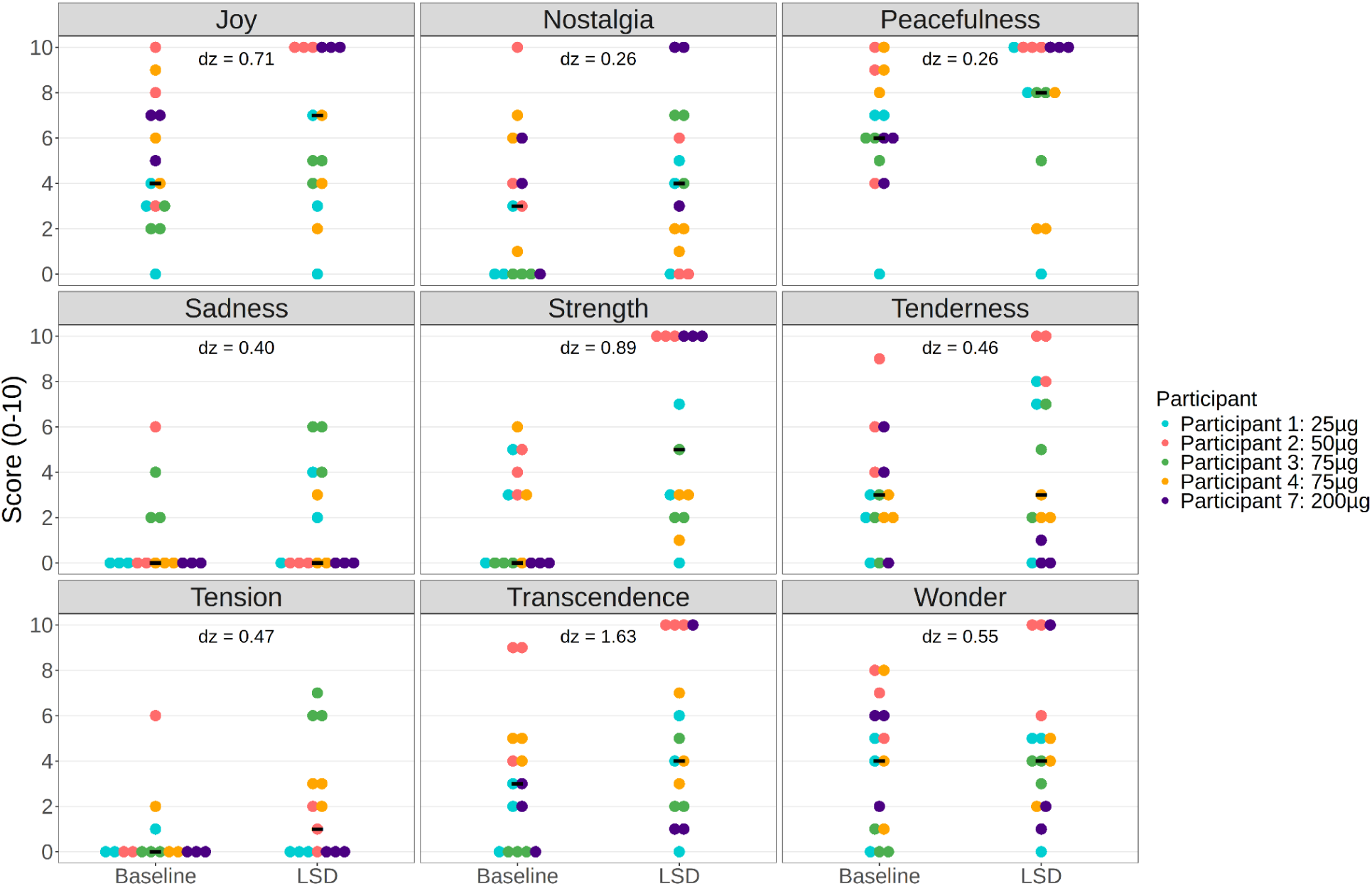
| Music-evoked emotional responses to LSD measured by Geneva Emotional Music Scale (GEMS). Music-evoked emotions that were measured within-scan using the GEMS at baseline and after LSD administration. The figure shows nine emotional dimensions: Joy, Nostalgia, Peacefulness, Sadness, Strength, Tenderness, Tension, Transcendence, and Wonder. Scores range from 0-10 on each dimension. Individual participant responses are shown as colored dots, while black diamonds represent group means. Effect sizes between conditions are indicated by Cohen’s d (dz) values for each emotional dimension.

**Supplementary Figure S14.**
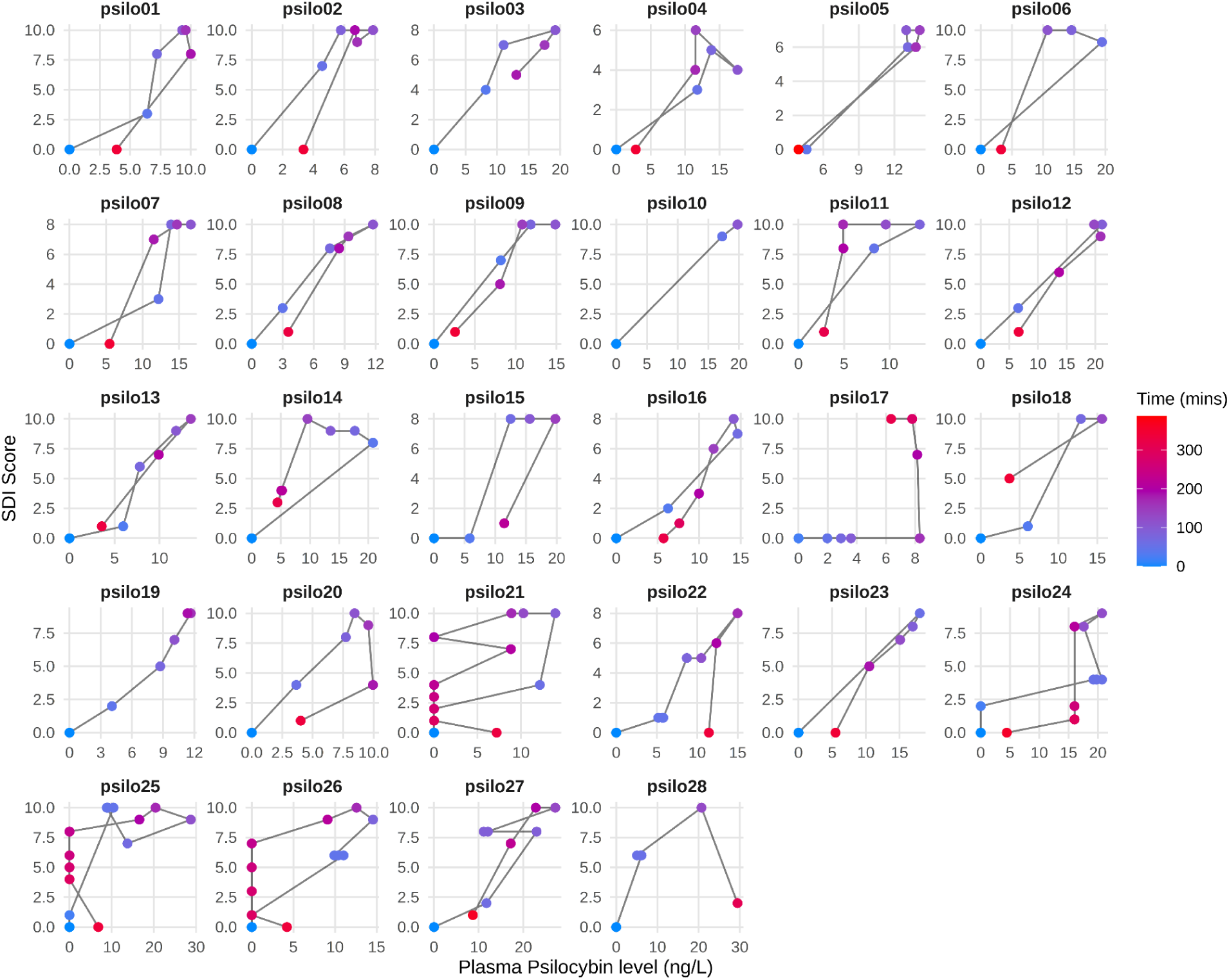
| Hysteresis curves showing the relation between plasma psilocin concentration and subjective drug intensity. Individual hysteresis curves for each participant (n = 28) showing the relation between plasma psilocin level (μg/L) and Subjective Drug Intensity (SDI) scores over time. Each point represents a measurement timepoint, coloured by time since psilocybin administration (minutes). Black lines connect sequential measurements, revealing the temporal trajectory of the subjective experience relative to plasma drug levels. The lack of consistent hysteresis pattern indicates no temporal dissociation between pharmacokinetics and pharmacodynamics.

**Supplementary Figure S15.**
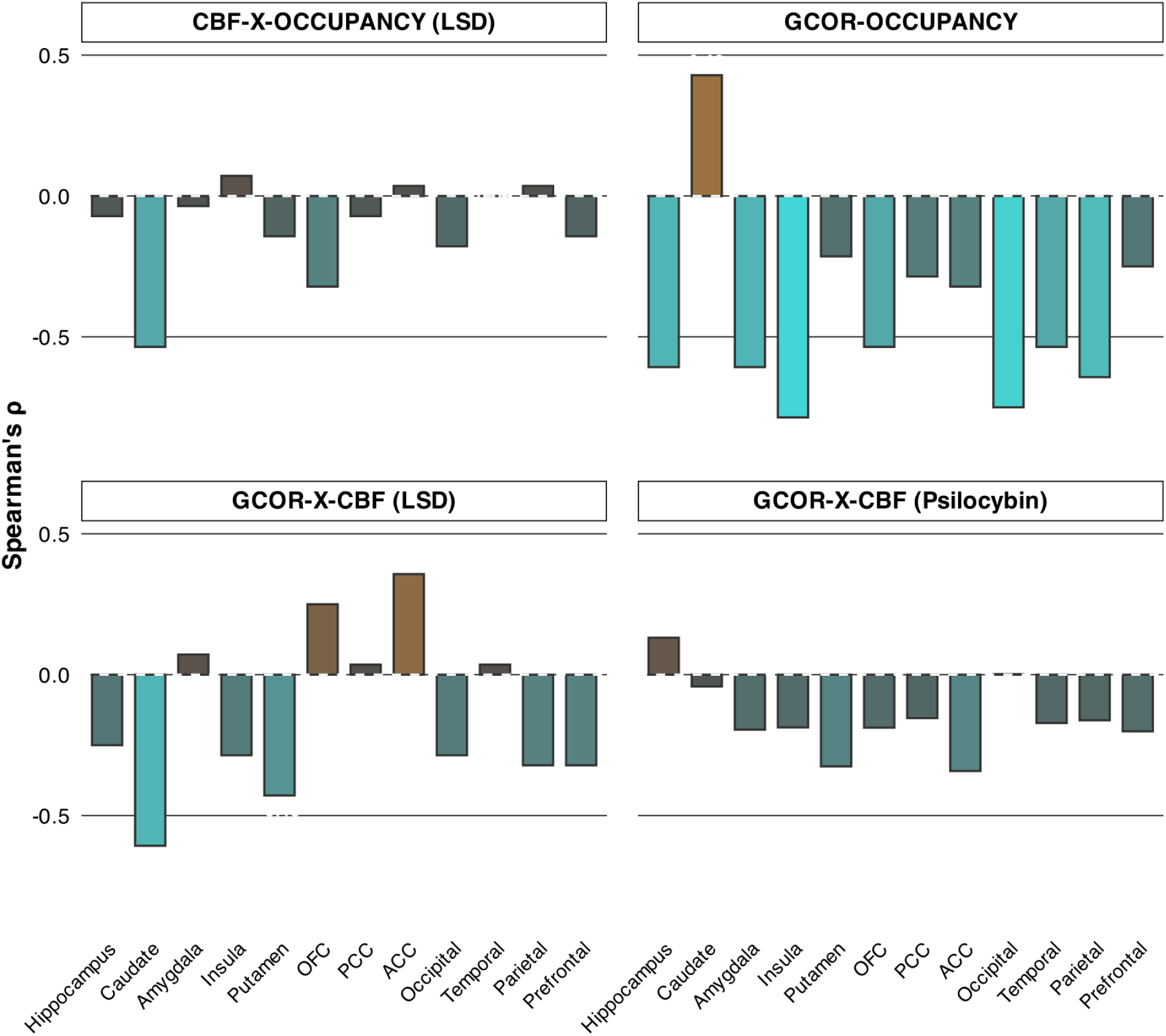
| Regional correlations between cerebral blood flow, global correlation, and receptor occupancy. Bar plots show Spearman’s rank correlations (ρ) for each brain region. Top left: Regional CBF changes versus neocortex 5-HT2A receptor occupancy in LSD. Top right: Regional GCOR changes versus neocortex occupancy. Bottom left: Regional GCOR versus CBF changes in LSD. Bottom right: Regional GCOR versus CBF changes in psilocybin.

**Supplementary Table S1.** | Participant characteristics and PET scan parameters across study phases. Demographics, physical characteristics, and study parameters for participants in baseline (n=11), peak LSD (n=7), and late LSD (n=2) scanning sessions. Data are presented as mean ± standard deviation with [minimum-maximum] ranges. Measures include age (years), sex distribution (Male/Female), weight (kg), height (cm), radiotracer parameters (injected dose in MBq and mass in µg), and number of previous psychedelic uses. *Only means are presented in “Late Scan” column to avoid reporting single-subject demographic information.

| Measure | Baseline | Peak Scan | Late Scan |
| --- | --- | --- | --- |
| n | 11 | 7 | 2 |
| Age | 31 ± 9[23-55] | 29 ± 6[23-41] | 28* |
| Sex (M/F) | 9 M / 2 F | 6 M / 1 F | 1 M / 1 F |
| Weight (kg) | 81 ± 12[63-106] | 84 ± 14[63-106] | 68* |
| Height (cm) | 184 ± 9[167-195] | 184 ± 8[170-192] | 175* |
| Injected dose (MBq) | 361 ± 117[147-501] | 334 ± 81[207-427] | 417, 436 |
| Injected mass (µg) | 0.21 ± 0.16[0.033-0.63] | 0.20 ± 0.11 [0.02-0.31] | 0.31, 0.38 |
| Previous psychedelic uses | 18 ± 43[0-146] | 8 ± 9[0-20] | 0, 1 |

**Supplementary Table S2.** | Regional cerebral blood flow (ml/100g/min), internal carotid artery (ICA) diameter (mm), and blood flow (ml/min) measures at baseline and after LSD administration. Values are presented as means (µ) with standard deviations (σ), medians (M), and ranges [,], along with numerical (Δ) and percentage (Δ%) changes, t-statistics, p-values (uncorrected and step-down corrected), and effect sizes (Cohen’s dz).

| MEASURE | Baseline |  |  | LSD |  |  | Change |  | Statistics |  |  |  |
| --- | --- | --- | --- | --- | --- | --- | --- | --- | --- | --- | --- | --- |
| | $\mu \pm \sigma$ | M | range | $\mu \pm \sigma$ | M | range | $\Delta$ | $\Delta\%$ | t-stat | p-value | p-stepdown | cohen's dz |
| **gCBF** | **49.2 ± 10.4** | **50.8** | **29.1 - 59.0** | **58.7 ± 10.9** | **57.1** | **45.6 - 77.4** | **6.3** | **19.3** | **3.16** | **0.020** | **NA** | **1.20** |
| Prefrontal | 53.7 ± 11.7 | 56.4 | 31.4 - 64.5 | 63.3 ± 11.4 | 60.1 | 49.4 - 83.5 | 3.7 | 18.0 | 2.85 | 0.029 | 0.125 | 1.08 |
| Parietal | 54.0 ± 11.4 | 54.8 | 31.9 - 67.5 | 63.8 ± 11.0 | 64.6 | 51.7 - 82.2 | 9.8 | 18.0 | 2.65 | 0.038 | 0.125 | 1.00 |
| Temporal | 49.9 ± 10.3 | 52.0 | 31.5 - 60.8 | 59.0 ± 12.7 | 55.3 | 42.5 - 83.1 | 3.4 | 18.4 | 2.21 | 0.069 | 0.125 | 0.84 |
| Occipital | 46.2 ± 9.8 | 47.3 | 28.5 - 56.2 | 58.4 ± 10.5 | 60.7 | 44.1 - 70.8 | 13.3 | 26.4 | 4.89 | 0.003 | 0.016 | 1.85 |
| OFC | 48.6 ± 11.0 | 52.8 | 29.4 - 59.3 | 55.8 ± 10.6 | 59.0 | 36.9 - 70.7 | 6.1 | 14.9 | 1.68 | 0.144 | 0.203 | 0.63 |
| ACC | 57.5 ± 13.3 | 64.1 | 35.1 - 68.8 | 66.9 ± 13.6 | 65.7 | 52.8 - 92.3 | 1.6 | 16.3 | 2.20 | 0.070 | 0.109 | 0.83 |
| PCC | 63.3 ± 13.9 | 64.3 | 40.3 - 79.3 | 74.2 ± 17.0 | 76.9 | 51.5 - 105.1 | 12.7 | 17.3 | 3.19 | 0.019 | 0.094 | 1.21 |
| Insula | 56.6 ± 11.8 | 64.4 | 37.0 - 66.6 | 67.9 ± 14.9 | 65.0 | 49.3 - 93.1 | 0.6 | 20.1 | 3.04 | 0.023 | 0.109 | 1.15 |
| Hippocampus | 44.3 ± 9.5 | 44.1 | 26.7 - 57.5 | 51.0 ± 12.7 | 47.1 | 39.0 - 74.1 | 3.0 | 15.1 | 2.34 | 0.058 | 0.141 | 0.88 |
| Amygdala | 39.5 ± 9.0 | 39.8 | 25.1 - 49.1 | 43.8 ± 14.5 | 37.3 | 33.0 - 73.4 | -2.4 | 10.9 | 0.95 | 0.380 | 0.406 | 0.36 |
| Thalamus | 40.9 ± 8.8 | 40.3 | 25.7 - 51.2 | 48.8 ± 6.7 | 50.4 | 39.1 - 55.7 | 10.2 | 19.4 | 2.76 | 0.033 | 0.141 | 1.04 |
| Caudate | 38.9 ± 11.1 | 41.0 | 20.6 - 52.8 | 46.5 ± 7.2 | 44.6 | 36.7 - 57.7 | 3.6 | 19.5 | 2.26 | 0.064 | 0.141 | 0.86 |
| Putamen | 43.3 ± 8.9 | 45.6 | 29.7 - 52.5 | 53.4 ± 11.2 | 50.3 | 37.7 - 72.6 | 4.8 | 23.2 | 2.71 | 0.035 | 0.125 | 1.03 |
| **ICA Diameter (mm)** | **4.5 ± 0.4** | **4.4** | **3.9 - 5.1** | **4.3 ± 0.6** | **4.4** | **3.3 - 5.0** | **0.0** | **-3.5** | **1.00** | **0.356** | **NA** | **-0.38** |
| **ICA Flow (ml/min)** | **210.1 ± 62.3** | **238.5** | **92.6 - 272.1** | **268.9 ± 51.9** | **270.7** | **221.3 - 369.1** | **32.2** | **28.0** | **-3.43** | **0.014** | **NA** | **1.30** |

## Additional Supplementary Tables

The following Supplementary Tables are available as Excel workbooks.

**Supplementary Table S3 | Global correlation (GCOR) changes following LSD administration.** Regional changes in global functional connectivity organized by brain region and network. The table includes baseline and LSD condition means, differences, t-statistics, effect sizes (Cohen’s dz), p values and confidence intervals for each brain region analyzed. Regions sorted by network membership according to the Schaefer 200-parcel atlas plus 32 subcortical regions from the Tian S3 subcortical atlas. Sheet 1 contains results without global signal regression and Sheet 2 contains results following global signal regression.

**Supplementary Table S4 | Functional MRI results.** Detailed results from analyses of network-level functional connectivity measures, including static and dynamic network connectivity, sample entropy at multiple scales, normalized spatial complexity, and global brain organization metrics (modularity, small-worldness, etc.). For each metric, the table presents t-statistics, p values, confidence intervals, effect sizes (Cohen’s dz), and uncorrected p-values. Sheet 1 contains results without global signal regression and Sheet 2 contains results following global signal regression.

**Supplementary Table S5 | AAL3 regions of interest.** Description of how we combined regions from the Automated Anatomical Labeling 3 (AAL3) atlas into 13 broader anatomical regions of interest (ROIs). Each row represents one of these ROIs, with the corresponding AAL3 subregions that were combined to create it. All regions include both left and right hemispheric components.

